# Accounting for place: confounding via geography obscures polygenic evidence on mental health and environmental exposures in the UK Biobank

**DOI:** 10.1101/2025.09.04.25335104

**Authors:** Zoe E. Reed, Tim T. Morris, Oliver S.P. Davis, George Davey Smith, Marcus R. Munafò, Gareth J. Griffith

## Abstract

Clustering of common causes by geography (confounding via geography) can induce bias if ignored. Analyses using polygenic indicators (PGI) for mental health traits are traditionally adjusted for genetic principal components (PCs), but not local area. In this study, we investigate whether accounting for geography more explicitly helps ameliorate such confounding. Using UK Biobank data (N=209,391 to 293,851), we construct PGIs across a range of p-value thresholds and contrast single-level regression with multilevel Mundlak models for the relationship between genetically predicted mental health and greenspace, whilst adjusting for 25 genetic PCs. Our formulation decomposes single level estimates into within-area, between-area and contextual estimates of mental health PGIs on two greenspace measures.

Single-level models for the least filtered PGIs find genetically predicted depression and schizophrenia associate with lower greenspace, while genetically predicted wellbeing associates with higher greenspace. However stricter p-value selection on PGI exposures results in estimates largely attenuating and, for schizophrenia, changing sign entirely. Within-area estimates of less-filtered PGIs align more closely with strictly filtered PGIs. Differences appear driven by between-area effects, with single-level models underperforming particularly in the most urbanised areas, and National Parks. We advocate for investigating the inclusion of local context as a routine sensitivity analysis for genetic epidemiologists working with spatially structured populations.

## Introduction

The observational association between geographical environment and mental health outcomes is well established. In 1939 Faris and Dunham (Faris & Dunham, 1939) observed a “spatial ecology” where greater rates of schizophrenia were observed in the innermost locations of urban Chicago. In the 86 years since this seminal work, causal understanding of these associations has remained limited. Largely opinions have been split into two camps; those arguing the association is a consequence of an urban environment causally acting on individuals; and others arguing that individuals predisposed to psychosis cluster in those environments due to behaviour-related migration (e.g. Giggs, 1975; Gudgin, 1975). The root of this debate likely lies in a phenomenon we term confounding via geography. Confounding via geography may bias any study investigating contextually clustered exposures or outcomes. In essence, exposures which are correlated within geographical contexts can open backdoor paths with other confounds due to the consistency (and thus non-independence) of observations within areas (see Box 1 for a more detailed explanation).

In an effort to unpick causality, contemporary researchers have increasingly supplemented phenotypic analyses of the psychiatric-environmental relationship with genetically predicted mental health exposures to control for an individual’s predisposition to a given mental health trait (Colodro-Conde et al., 2018; Maxwell et al., 2021). Moreover, whilst the association between schizophrenia and urbanicity is now well known, contemporary psychiatric spatial ecologies have found popularity in establishing an observational association of greenspace exposure with both better mental health and increased wellbeing (Engemann et al., 2019; Feng & Astell-Burt, 2017; Liu et al., 2024; Marcham & Ellett, 2024; Zare Sakhvidi et al., 2023; Zhang et al., 2020).

Here, we use the greenspace and mental health case as a motivating example to investigate whether confounding via geography may bias genetic epidemiological associations.

### Box 1

**Confounding via Geography in Epidemiological Studies**.

*In 1970, Waldo Tobler popularised the idea that it is the norm rather than the exception that individuals residing in the same geographical location are more similar than individuals from geographically distant areas (Tobler, 1970)*.

*This geographical clustering of observations typically undermines efforts at causal inference through two principal mechanisms: confounding, and interference*^*1*^. *For the purposes of this paper, we focus on geographical confounding. Confounding via geography can occur where common causes are clustered by geographical context. Whilst geography may not act as a true cause in such cases, it represents a meaningful context in everyday life at which such causes cluster – via, for instance, local policy decisions or pollutant* exposures.

*As with any causal inference analysis, omitting common causes risks inducing a statistical relationship between otherwise unrelated variables. Furthermore, geographical clustering may violate assumptions of the independence of modelled residuals, as such residuals often cluster within geographical contexts – leading to deflated standard errors and greater type 1 error risk (Pickett & Pearl, 2001; Subramanian et al., 2009*).

*Problematically, for many geographically structured epidemiological analyses, causally-relevant geographical confounds are often unmeasured, and functionally often difficult or impossible to capture accurately. Imagine, for instance, trying to adjust for every causally relevant reason that residing in London differs from residing in Berlin. The causal complexity and inter-dependence of the historical, political-economic, environmental, and socio-demographic characteristics likely renders any proposed adjustment set inadequate*.

*Whilst such confounding via geography presents a risk to causal inference if ignored, when appropriately addressed, integrating geographical context into analyses can offer researchers a mechanism for inferring about complex causal structures. Here we demonstrate the benefits of deploying a geographically informed methodology to simultaneously and explicitly model within-area processes and between-area processes*.

^1^*Geographical interference (sometimes termed dependence or spillover) presents a separate but related inferential problem, where the exposure in Area 1 affects outcomes in Area 2. If such interference exists and is not explicitly modelled then this likely violates the Stable Unit Treatment Value Assumption (SUTVA) (Angrist et al., 1996). A more complete discussion of spatial interference and consequences of SUTVA violations can be found elsewhere (Akbari et al., 2021)*.

Confounding via geography is a thorny issue for epidemiologists interested in the causes of geographical differences in psychiatric disorders, as both mental health outcomes (e.g., depressive symptoms (Chaix et al., 2006; Griffith & Jones, 2020)) and environmental measures (e.g., greenspace access (Mitchell & Popham, 2008)) exhibit strong geographical patterning in the UK. As outlined in Box 1, such patterning may induce correlations with other geographically patterned confounds sharing the same structure.

Considering our motivating example; we wish to know “Does mental health predispose individuals to move to greener or less green locations?”. Figure 1 demonstrates why considering a large-scale dataset to analyse this individual-level relationship may be problematic if we ignore geographical context. Namely, if we treat individuals as independent, we may misattribute variance truly explained by geographically structured confounds (e.g., demographic differences, climatological exposures, or local policy contexts) to individual mental health exposures.

**Figure 1.**
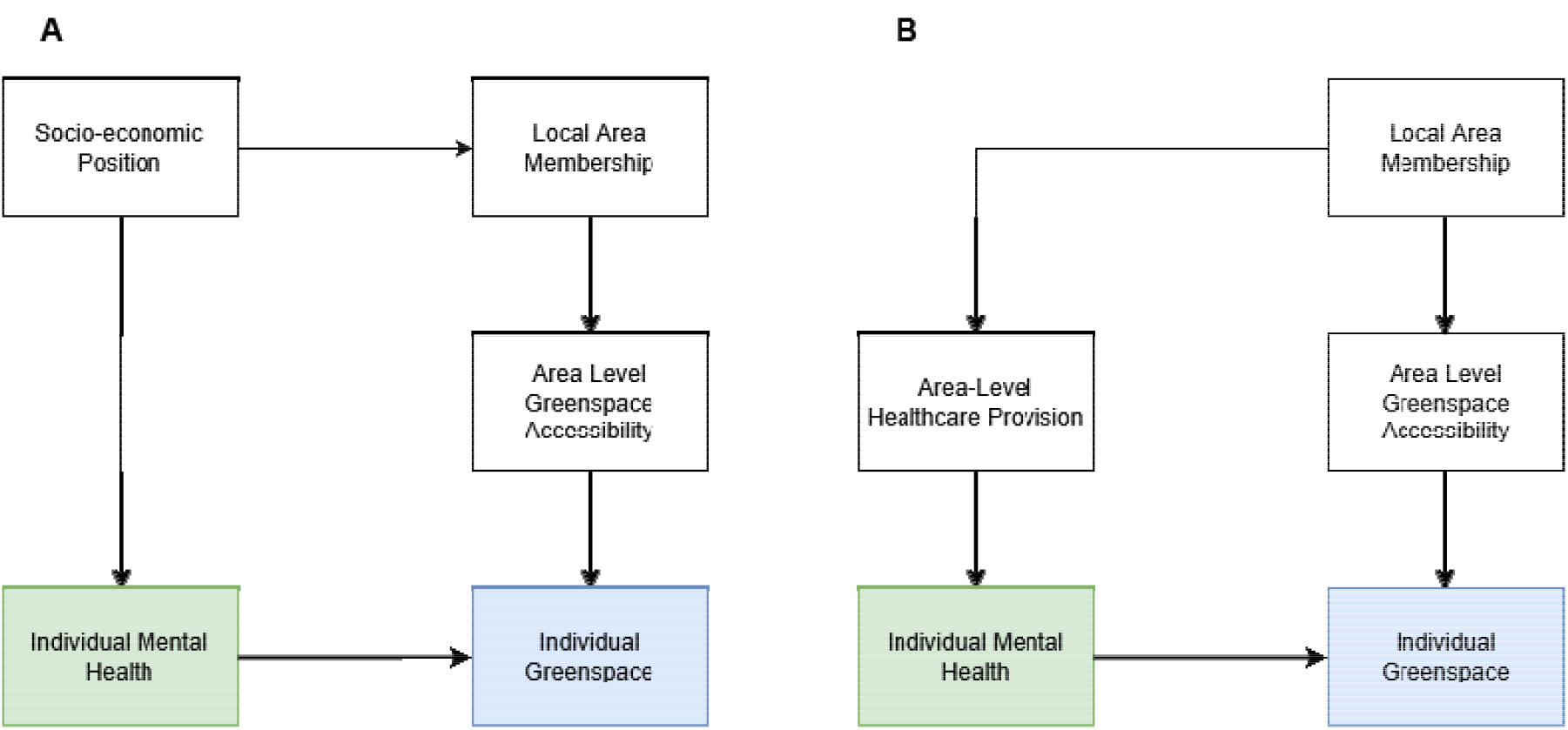
Directed Acyclic Graphs (DAGs) illustrating two conceptual models, with geographically patterned confounding structures between individual mental health exposures, and greenspace outcomes. 1A illustrates a backdoor path where socio-economic position is assumed to exert a causal influence on local area-membership, through migration, and 1B illustrates a scenario where a backdoor path is opened between individual level mental health and greenspace solely through the geographically informed local policy conditions.

Figure 1 illustrates some plausible instances of confounding via geography using Directed Acyclic Graphs (DAGs). In Figure 1A socio-economic position (SEP) causes individual mental health outcomes and area-membership (e.g., through house price constraints and selective migration), which opens a backdoor path through SEP. Figure 1B demonstrates a similar scenario, where the backdoor path is explicitly via area-membership (through local policy conditions relating to healthcare provision). If such geographical structure, and structured confounds are not explicitly modelled by researchers, then the variance associated with geography will be distributed over the structural levels within the model, risking Type 1 error, and deflated standard errors (Tranmer & Steel, 2001).

The causal structures described in Figure 1 become more complex when we consider genetically informed exposures (specifically polygenic indices, PGIs, derived from genome-wide association studies, GWAS). Genetically informed exposures are known to exhibit spatial structure (Abdellaoui et al., 2022; Haworth & Davis, 2014); however, the degree and nature of spatial structure in the population are largely unknown. As outlined in Box 1, area-membership cannot directly cause individual PGI; however both the trait a PGI relates to and area-membership may share a common cause via measured or unmeasured confounders such as those above. This common cause may then induce geographical structure per Figure 1A.

Figure 2 provides DAGs for a within-area and multiple area formulation of the influence of mental health PGI on individual greenspace. For illustrative purposes we have displayed parental SEP as a geographically structured confound but C may represent any geographically clustered (likely unmeasured) confound. Figure 2A illustrates this in a single-area context without selective migration or population structure, such that parental SEP and individual mental health PGI are independent, and we need not condition on parental SEP. Whilst convenient, Fig 2A will be an oversimplification in all real-world applications due to the geographical structure of most data generating sampling frames.

**Figure 2.**
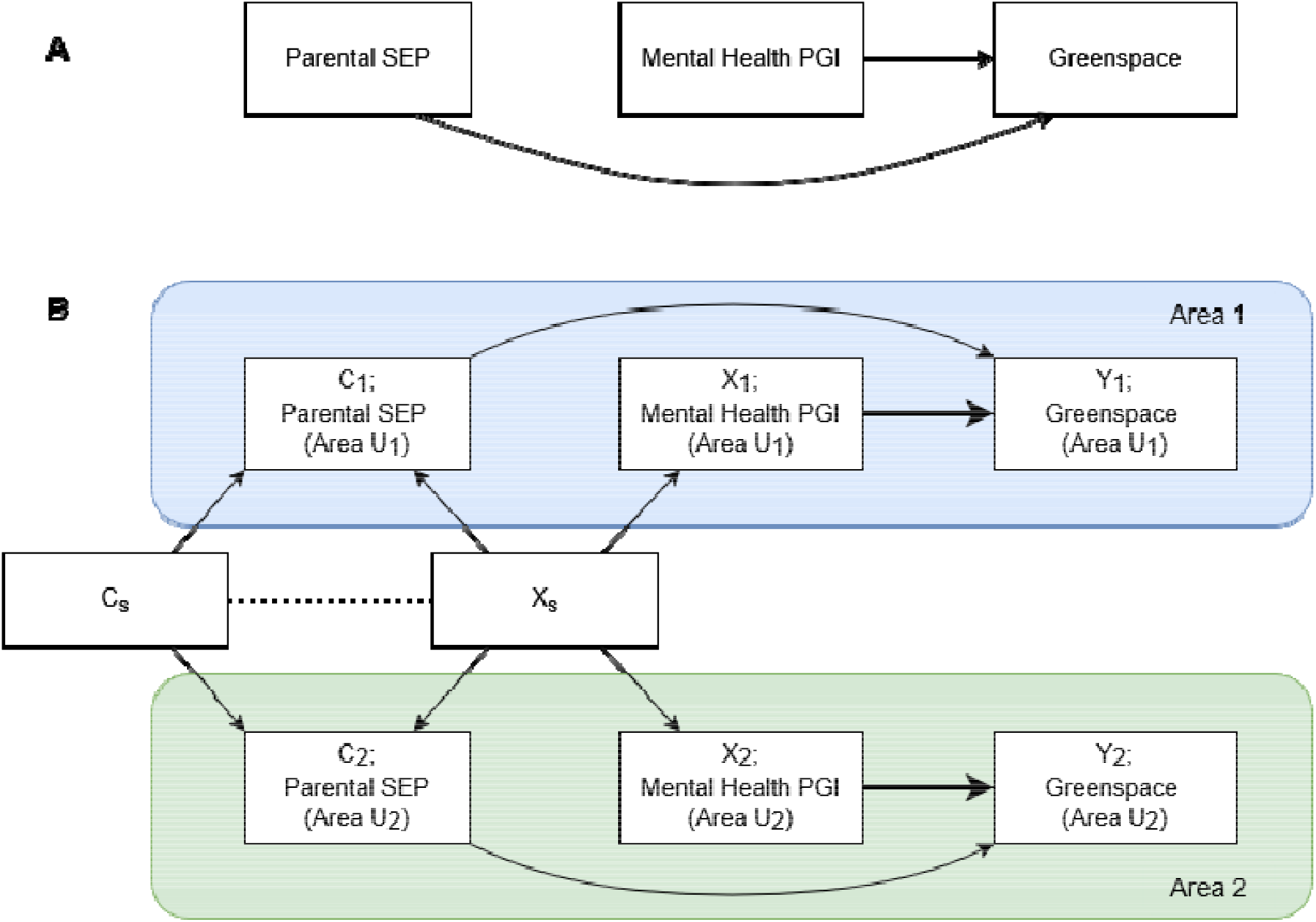
DAGs illustrating a backdoor path between mental health polygenic indices (PGIs) and greenspace via the geographical patterning of the exposure and confounders when considering multiple geographical contexts. Figure 2A illustrates a single level formulation, and Figure 2B demonstrates the backdoor paths opened by implicitly considering the geographical structure of the PGI exposure (X_s_) and confounds (C_s_) in a dyadic two-area example.

Following DAG convention from Papadogeorgou and Samanta (Papadogeorgou & Samanta, 2024), in Figure 2B we present a multiple area model where there are two hypothetical areas, U_1_ and U_2_, presented in the upper and lower halves of the DAG. As there are multiple area contexts – we explicitly incorporate nodes for the geographical structure of the exposure and confounders, indexed by a subscript S. The geographical structure of parental SEP is given by C_S_ and the geographical structure of the polygenic index is given by X_S_. Importantly, X_S_ captures the contemporary complex consequences of population stratification and migration events. In Figure 2B, parental SEP does not cause individual level PGI, however they share a common cause via the processes which give rise to the geographical structure X_S_, which may or may not concord with the geographical structure of C_S_ (indicated via a dashed line).

Using graph theory, we can see that if we do not perfectly condition on the geographical structure of the PGI (X_S_), there is an open backdoor path from X to Y through X_S_ and C. The degree to which these X_S_ and C_S_ are correlated (indicated by a dashed line) for any given unobserved C, will determine the degree to which other geographical confounds (such as those hypothesised in Figure 1) may also bias our observed relationship between X and Y, via C_S_.

Where a PGI is suspected to be confounded via geography (for instance via population structure), modelling geographical context allows researchers to interrogate the degree to which the population association reflects within-area and between-area signal.

To close the paths through X_S_ we must both account for the similarity between observations within an area on unmeasured confounders (C_S_), and account for within-area similarity in exposure (X_S_). To satisfy such criteria, researchers may additionally adjust for area-level means for PGI exposures.

The same approach is often carried out in genetic studies using family designs to remove between-family differences, by taking the departure of an individual’s genotype from the mean genotype for each family unit (Howe et al., 2022). Here we adopt the same framework but use geographical context instead of a family unit. However, we go further, explicitly modelling this variance, rather than including dummy terms for each area-mean, treating the geographical variance as of substantive interest rather than a nuisance to be removed. Explicitly interrogating this geographically structured variance, requires an operationalisation of a multilevel random effects framework proposed by Mundlak (Mundlak, 1978).

In Mundlak’s formulation, the cluster-mean exposure is fit alongside the individual level exposure, as per a traditional between-within decomposition. However, in the Mundlak formulation, the cluster-mean exposure is not subtracted from the individual level observation. This results in the area-mean term representing what is termed a *contextual* effect estimate. A contextual effect estimates the effect of moving one individual with fixed individual level characteristics (including PGI) to an area with a different-mean PGI. The Mundlak formulation also means that the effect of any geographically clustered similarities between areas (C_S_) which are consistent with the geographically structured exposure (X_s_) will be captured by the contextual effect estimate. This formulation then, provides us the capacity to learn about the gene-environment correlations that exist in our data, via this contextual effect estimate of the effect of moving between areas with different mean PGI (for more detailed discussion see Supporting Information Section 4).

Single-level models necessarily assume that between- and within-area effects are identical, thus assuming that this contextual effect is zero. Here we specify that for our given example, within- and between-effects are conceptually distinct. For our model, a within-area effect captures the average greenspace response for a unit change in mental health PGI, *within the average neighbourhood*, having accounted for between-neighbourhood differences. The contextual effect gives the effect of an individual moving from one neighbourhood to another where the mean PGI is 1 unit higher, holding their individual characteristics constant. We argue that genetic epidemiologists are often interested in the within-area effects, as between area differences as a consequence of area-level exposures are not likely to reflect the true causal pathway of interest. If the between- and within-area estimates are zero (as assumed in the traditional single level model) then the contextual effect estimate should simply be zero in our modelling framework. If, however, the contextual effect is non-zero, then it is likely that the within-area estimate is more likely capturing the effect estimate of interest.

We demonstrate the framework above to evaluate the relationship between PGI for three mental health traits (depression, wellbeing and schizophrenia) and two reported phenotypic greenspace outcomes in UK Biobank, adjusting for the first 25 genetic principal components of population structure in all analyses. We estimate within-area and contextual effect estimates of mental health PGI at a number of p-value thresholds on greenspace outcomes in each of our models. The intuition here is that if there is greater concordance between the within- and single-level results when a PGI is more strictly filtered, then this mechanism may be included as a sensitivity analysis for common genetically informed associations, and genetic epidemiologists ought to routinely consider within-area estimates as a test for unmeasured geographical structure in the data (similar to the Hausman test (Hausman, 1978)).

Given the substantive and statistical distinction between a contextual and within-area effect, we are interested in the degree to which single level effects may be attributing confounding via geography to an individual-level effect. We can test the validity of the assumption of zero contextual effect via estimating it explicitly through the Mundlak formulation (algebraic specification in Supporting Information Section 4). If non-zero contextual effects are found, this implies the between effect differs from the within-area effect, indicating greater chance of confounding via geography. We also wish to learn whether we can recover the more robustly filtered genetic signal via the within-area estimator – which would indicate that genetic epidemiologists might consider routinely deploying a clustering mechanism similar to the Hausman test.

### Research Questions

To evaluate this, we present the following three research questions:

1. Is there evidence that individuals with a higher genetic propensity to depression or schizophrenia, and lower genetic propensity for wellbeing reside in less green locations?
2. Is this within-area effect of this mental health PGI on greenspace consistent with the single-level estimate?
3. Do more strictly filtered PGI have greater concordance between single-level and within-area estimates for effects on greenspace?

We compare results across two common parameterisations of greenspace, and make recommendations for future researchers interested in the effects of geographically or otherwise contextually clustered genetic exposures.

## Results

We describe the phenotypic sample characteristics for our analytic sample in the UK Biobank in Table 1. We then linearly regress standardised greenspace outcomes (both NDVI and greenspace) on each of the phenotypic exposures (Supplementary Table S1).

**Table 1.** UK Biobank sample characteristics.

|  | **N with data available** | **Mean (SD) or prevalence** |
| --- | --- | --- |
| **Age** | 336,902 | 56.87 (8.0) |
| **Sex (Female)** | 336,902 | 53.8% |
| **NDVI** | 209,391 | 0.11 (0.14) |
| **Percentage greenspace** | 293,851 | 36.9 (23.6) |
| **Depression diagnosis** | 336,902 | 14.4% |
| **MHQ depression** | 79,922 | 26.8% |
| **Wellbeing (1 to 6)** | 109,912 | 4.6 (0.8) |
| **Schizophrenia diagnosis** | 336,158 | 0.2% |

*SD=Standard Deviation, NDVI=Normalised Difference Vegetation, MHQ=Mental Health Questionnaire*

### Phenotypic analyses of greenspace and NDVI on mental health phenotypes

We find strong evidence of negative associations between greenspace and each of depression diagnosis (−0.06, 95%CI −0.08 to −0.06), MHQ depression (−0.03, 95%CI −0.05 to −0.01), and schizophrenia (−0.32, 95%CI −0.40 to −0.25). There is also clear evidence of a positive association between wellbeing and the greenspace outcome (0.06, 95%CI 0.05 to 0.07).

There was little evidence of phenotypic associations between NDVI and depression diagnosis (−0.005, 95CI −0.02 to 0.007) or schizophrenia (0.04, 95%CI −0.05 to 0.13). There was evidence of a negative association with MHQ depression (−0.02, 95%CI −0.04 to −0.004), as well as with wellbeing (−0.01, 95CI −0.02 to −0.0003) which had a different direction of effect to results for greenspace.

### Regression of greenspace and NDVI on PGIs for depression, wellbeing and schizophrenia

To explore the causality underpinning the phenotypic relationships we regress standardised greenspace on PGI exposures for three traits, each with single nucleotide polymorphisms (SNPs) selected at a range of p-value thresholds. We specify within-area and single-level estimators (algebraic derivation is given in Supporting Information Section 4) to explore the degree to which a within-area estimator can recover the effect estimate of a more strictly p-value filtered genetic signal from the noisier signal of a less strictly p-value filtered index. Supplementary Tables S2 and S3 present results from standard single level models which combine within- and between-components, which are assumed equivalent, preventing insight into how between- and within-effects may differ.

Both outcomes exhibit a high degree of clustering, indicated by neighbourhood-level variance components. Greenspace has a level-2 Variance Partition Coefficient (VPC) of 0.48, whereas NDVI has a VPC of 0.79, implying that 48% and 79% of the unexplained variation in land use-derived greenspace and satellite derived greenspace can be explained by the neighbourhood^1^ the individual resides in. In a 2-level formulation, the VPC also implies that pairwise selections of individuals within the same area will (on average) be correlated 0.48 and 0.79 for greenspace and NDVI respectively.

Results from multilevel models are presented below in Figures 3 and 4 for greenspace and NDVI respectively. Full results for the within-area effects are presented in Supplementary Tables S4 and S5, and full results for the contextual effects are presented in Supplementary Tables S6 and S7, for greenspace and NDVI respectively. Supplementary Figures S4 and S5 present the estimated contextual effect estimate alongside single-level and within-area estimates.

**Figure 3.**
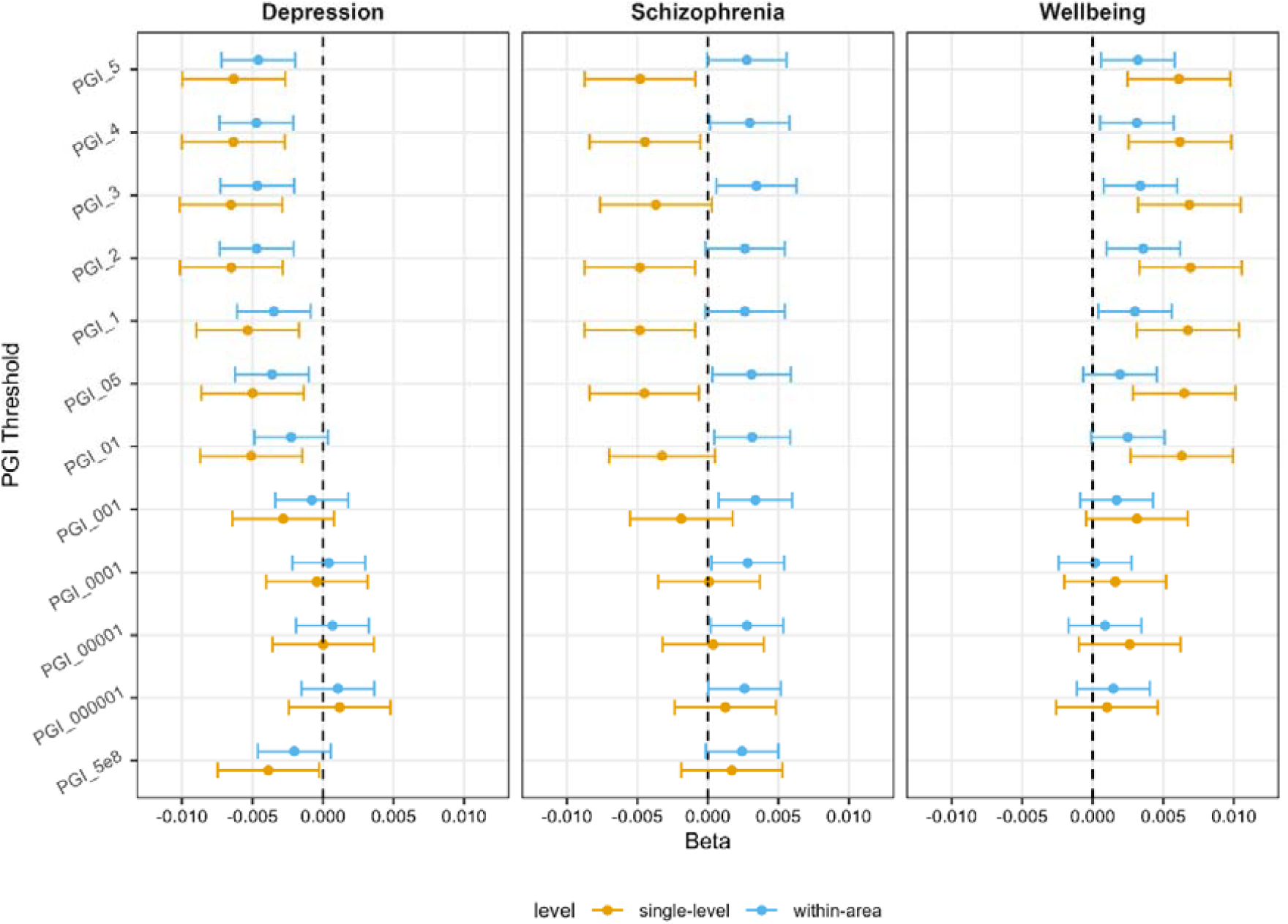
Estimates for single-level and within-area effects of mental health polygenic scores and the standardised percentage greenspace outcome. Single-level effects are taken from single level linear models, and the within-area effects are taken from Mundlak models. PGI_5 through PGI_5e8 denote polygenic indices constructed with SNP selection filtered at p<0.5, p<0.4, p<0.3, p<0.2, p<0.1, p<0.05, p<0.01, p<0.001, p<0.0001, p<0.00001, p<0.000001 and p< 5×10^−8^ in descending order down the Y axis.

**Figure 4.**
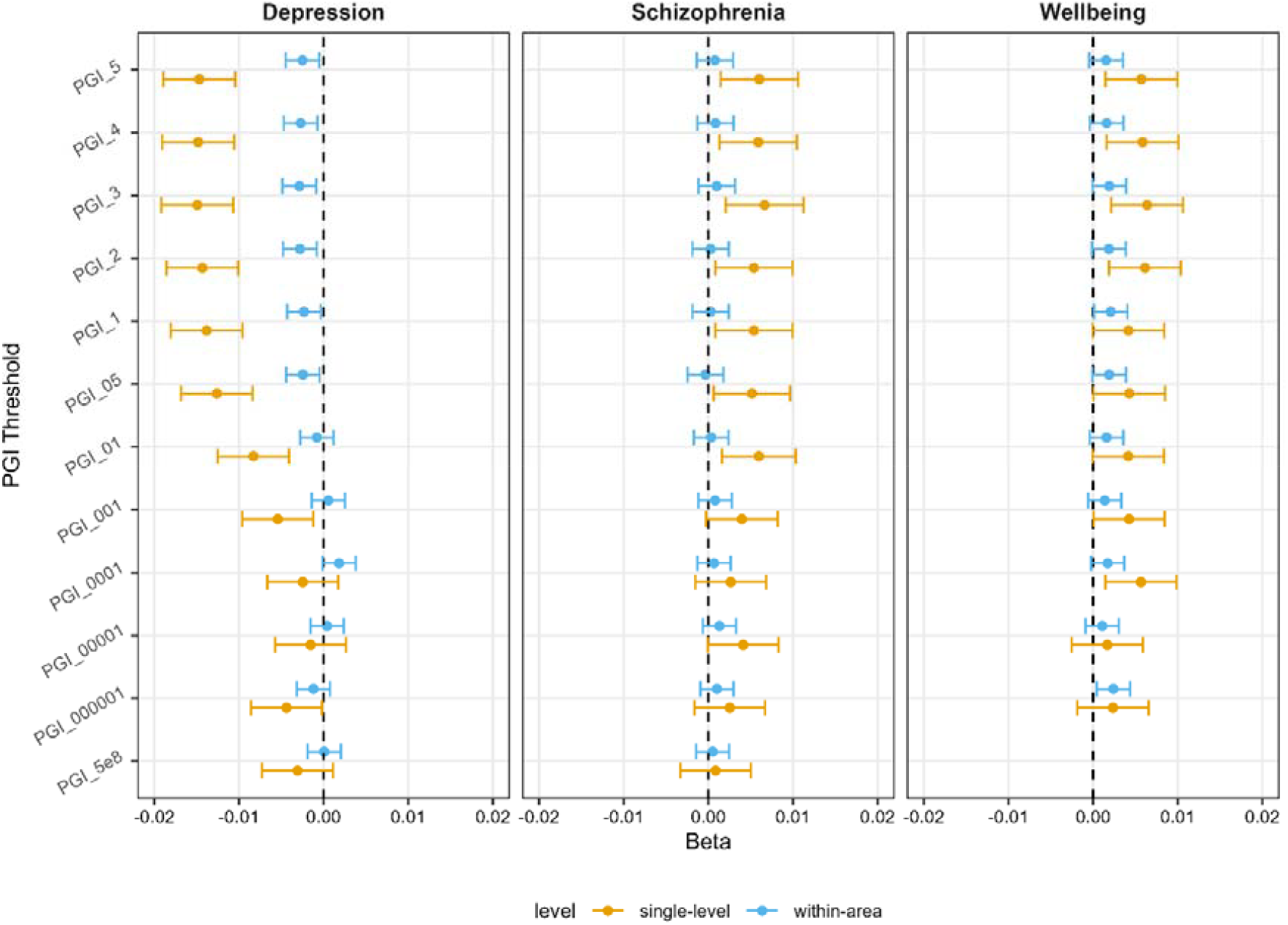
Estimates for single-level and within-area effects of mental health polygenic indices and standardised percentage NDVI. Single-level effects are from the single level linear models and the within-area effects are from the multilevel models. PGI_5 through PGI_5e8 denote polygenic indices constructed with SNP selection filtered at p<0.5, p<0.4, p<0.3, p<0.2, p<0.1, p<0.05, p<0.01, p<0.001, p<0.0001, p<0.00001, p<0.000001 and p< 5×10^−8^ in descending order down the Y axis. PGI = Polygenic Index.

Figure 3 shows that for PGI filtered for genome-wide significant hits (where included SNPs are anticipated to be more reliable) there is little difference between the within-area and the single-level estimate. Supplementary Figure S4 plots the contextual estimate alongside the estimates in Figure 3 to illustrate the relative magnitude of the contextual effect. There is some evidence from the linear formulation that genetically predicted depression (p-value 5e-8) is associated with lower percentage greenspace (−0.004, 95%CI −0.007 to −0.00023). We also see evidence that genetically predicted wellbeing (only at p-value thresholds >0.1) is associated with higher percentage greenspace (0.007, 95%CI 0.003 to 0.01). Finally, we see evidence that genetically predicted schizophrenia (only at p-value thresholds >0.05) is associated with lower percentage greenspace (−0.005, 95%CI −0.008 to −0.0006). At less strict p-value thresholds, we see larger discordance between the within-area and the single-level estimate. Schizophrenia results are particularly striking for the less strictly filtered PGI, suggesting that living in an area with higher average PGI is associated with *less* greenspace, but within a given area, those with higher schizophrenia PGI appear to be closer to *more* greenspace.

Supplementary Figure S4 illustrates that this is driven by the strong negative contextual effect of schizophrenia (e.g. schizophrenia PGI on land-use derived greenspace, p<0.5, (single-level: −0.005, 95% CI −0.009 to −0.0009, within: 0.003, 95% CI −0.00006 to 0.006, contextual: 0.03, 95% CI −0.07 to 0.01). Contextual estimates have far wider confidence intervals however, as the number of neighbourhoods is far smaller than the number of individuals. Across all outcomes, but particularly for schizophrenia and wellbeing, the within-area estimate for the less strictly filtered PGI recovers an effect estimate consistent with single-level estimator for more strictly filtered PGI.

### Regression of greenspace (NDVI) on PGI for depression, wellbeing and schizophrenia

Figure 5 presents the same exposures but predicting NDVI-derived greenness. There is again evidence of larger effect sizes in less strictly filtered PGI; however, consistent with the larger VPC for NDVI, these attenuate more strongly when adjusting for local context, and when filtering PGI more strongly. We see that at the most strictly filtered p-values, there is again little evidence of a relationship between genetically predicted mental health outcomes and NDVI-derived greenness.

**Figure 5.**
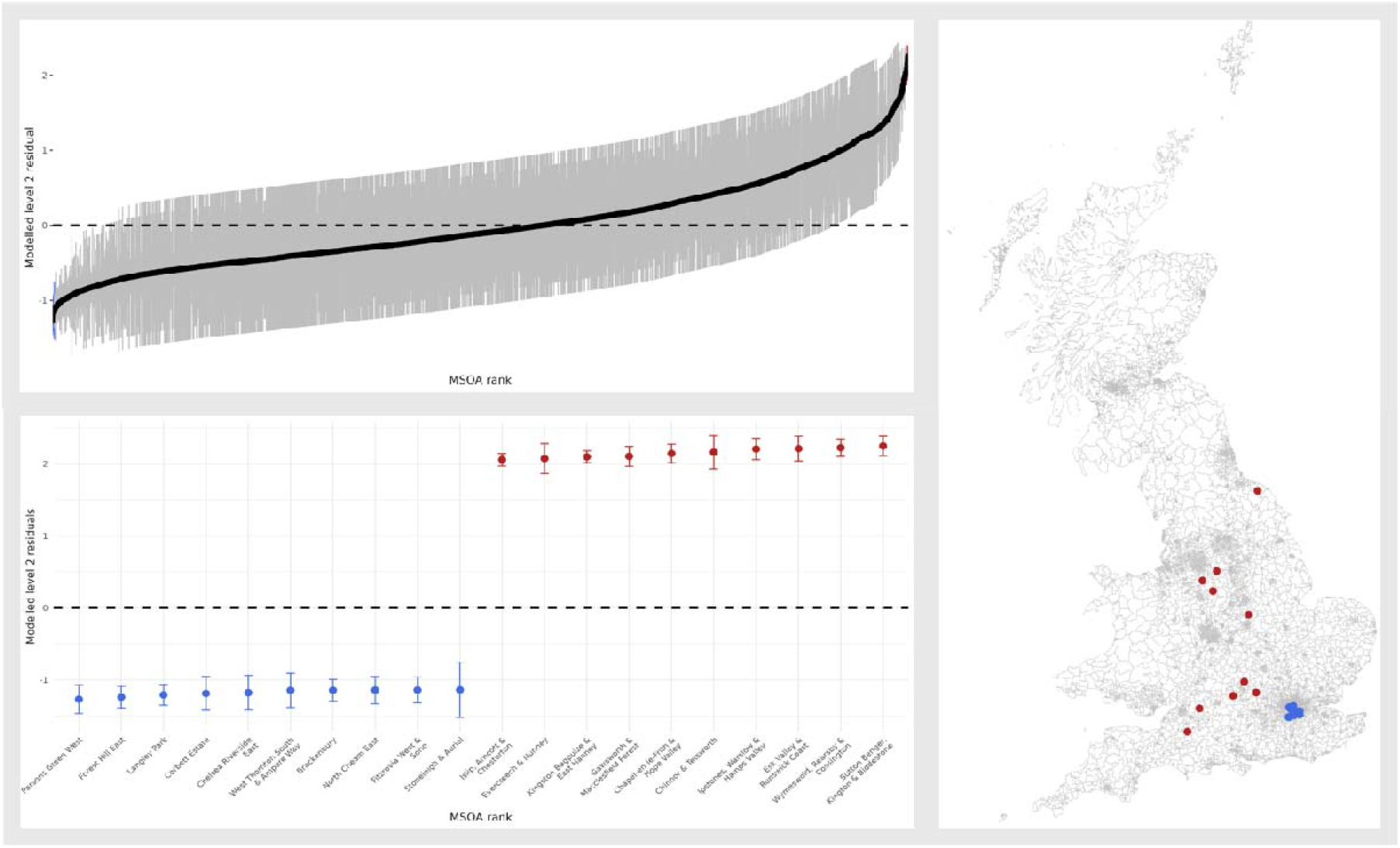
Ranked MSOA level residuals (top left) for the schizophrenia PGS (p< 5×10^−8^) and land-use derived greenspace model. Bottom left: Ranked MSOA residuals from Mundlak model filtered to solely display the top and bottom 10 MSOAs. MSOA labels are given on the X axis. Right: Mapped locations of the extreme MSOA residuals across the dataset, with highest residuals in red, and lowest residuals in blue.

Supplementary Figure S5 illustrates the contextual effect of NDVI on the same scales as the within-area effects above. The same confidence intervals are similarly broad for contextual estimates, but we again see evidence of large contextual effects, particularly for schizophrenia.

Figure 5 shows modelled neighbourhood residuals from our within-area Mundlak framework, taken from the strictly filtered schizophrenia-PGI greenspace model. It is worth reiterating at this point that all our models were adjusted for 25 genetic principal components, which are commonly included to adjust for population structure, but clearly do not adequately adjust for geographical structure. Extreme residuals indicate places where geographically naïve models underperform (particularly in areas with relatively extreme levels of greenspace, i.e., very rural or urban locations). Figure 5 illustrates that neighbourhoods with highest modelled MSOA residuals are in extremely rural areas, including national parks such as the Peak District, the Cotswolds and the North York Moors. Conversely, all of the 10 lowest modelled MSOA residuals are within London. Assuming conditional exchangeability between individuals in the North York Moors and London is likely incorrect, given structural and systematic differences between London and the North York Moors beyond that of greenspace or polygenic indicators. This may be a particularly thorny issue for studies like the UK Biobank, where recruitment centres were predominantly in heavily urbanised areas.

## Discussion

The phenotypic association of mental health exposures with greenspace is consistent with that found by previous research (Engemann et al., 2019; Feng & Astell-Burt, 2017; Liu et al., 2024; Marcham & Ellett, 2024; Zare Sakhvidi et al., 2023; Zhang et al., 2020). However, we find the strength of observed associations likely reflects confounding via geography. Genetically predicted mental health exposure estimates are attenuated compared with phenotypic exposures and attenuate further, in some cases changing the direction of effect, on more stringent SNP filtering in PGI, and appropriately accounting for geographical context. Using geographically sensitive approaches, we find, contradicting many previous studies, that individuals in the UK Biobank with a higher genetic propensity for schizophrenia or lower genetic propensity for depression are more likely to move to areas with more usable greenspace. Such a result may be explained by collider bias, where higher schizophrenia PGI is negatively associated with participation and higher greenspace is positively associated with participation (resulting in participants with high schizophrenia PGI being selected for higher greenspace).

We demonstrate that for certain outcomes we recover effects of more robustly filtered PGI estimates, from noisier polygenic indices with explicit consideration of geographical context. Given all our analyses adjust for 25 principal components of population structure, this suggests confounding operating via geographical structure that is not captured by genetic PCs. This is likely especially true of our study as greenspace is highly clustered, however, it is not unusual for researchers to deploy genetically informed methods for clustered exposures such as income, education or even air pollution (Hill et al., 2019; Okbay et al., 2022; Qiu et al., 2023).

The level-2 VPCs were very high across both outcomes. VPCs of 0.48 and 0.79 for land-use based percentage greenspace and land-satellite derived greenspace respectively, imply that 48% and 79% of the variance in the individual level observations can be explained by their MSOA membership. The VPC also implies that pairwise selections of individuals from within the same MSOA would be correlated on average to 0.48 and 0.79 for greenspace and NDVI respectively, further highlighting geographically structured non-independence.

Figure 5 highlights the least conditionally exchangeable MSOAs in the model of schizophrenia PGI (p filtered at 5×10^−8^). In a single level model, any differences between outcomes in central London, and national parks would be attributed solely to differences in PGI, conditional on covariates. Whilst this may be appropriate for some relationships, we suggest that researchers may need to interrogate this assumption more comprehensively in genetic epidemiological studies.

### Strengths and Limitations

Our results have two key analytical strengths. The first is that we are able to decompose a confounded estimate of a given PGI using the within-area formulation. We can see that this can cause substantial change to coefficients, particularly in the case of suspected noisy genetic signals (i.e., less strongly filtered PGI). We suggest that within-area estimands provide a more robust estimate of the putatively causal individual effect of interest, due to insulating against confounding via geography.

Secondly, due to partial pooling of our area level information, we do not exclude areas with low coverage, as these are shrunk towards the grand mean proportional to their robustness. Our data cover a wide range of geographical contexts (5,728 MSOAs for greenspace, and 4,896 for NDVI, see Supplementary Figure S4), implying that we are likely to have reasonable estimation of the contextual effects of PGI, but find that these are limited in power. This may be for a number of reasons, including further residual confounding within the area-mean terms, or the mis- or under-specification of geographical scale. Geographically informed models are highly sensitive to scale selection, for instance if confounding via geography truly occurred due to exposures consistent within governmental regions, not MSOAs, then the above model looking into MSOA-level confounding via geography may be overly adjusted (Griffith et al., 2021; Jones et al., 2015).

However, our study has several limitations. Firstly, we only capture the impact of geography in a hierarchical membership sense, where we model neighbourhood membership but not spatial adjacency within that membership which may lead to overstated precision (Wolf et al., 2021). More explicitly, our model cannot consider spatial contiguity, spatial networks, or spatial proximity beyond that within-neighbourhoods, such that if there are violations of SUTVA which relate to exposure to genetic indices of spatially proximal neighbours we may not capture them perfectly here.

Secondly, the UK Biobank is well known to be a selected population – healthier and wealthier than the age-adjusted UK population. This might induce artefactual associations between our exposures and outcomes if both relate to selection (Lu et al., 2024). Whilst we insulate against a certain degree of area-specific selection processes in our contextual formulation, the area-adjustment is agnostic to the reasons for the observed geographical structure. If such selection effects vary between assessment centres, this may bias associations for environmental variables and participation patterned phenotypes such as mental health.

The UK Biobank is particularly susceptible to selection bias when considering environmental exposures as recruitment took place in physical assessment centres, meaning geographical coverage does not enumerate all neighbourhoods in all constituent countries (see Supplementary Figures S2 and S3). This incomplete geographical coverage means when we are adjusting for “neighbourhood means” we are not truly adjusting for environmental characteristics of neighbourhoods, rather the means for observed individuals – which under selection is unlikely to represent true geographical structures.

### Implications

Disentangling the causal contributions of between- and within-area effects is of particular interest to genetic epidemiologists, as assuming between- and within-area equality makes separating gene-environment interaction (effect modification by geographical context) from gene-environment correlation (confounding via geography) extremely difficult. The degree of bias induced by confounding via geography is proportionate with the degree of clustering in the outcomes and exposures of interest (and respective confounds). Ongoing processes of social and spatial segregation, as well as spatially informed selection mechanisms mean that such biases are likely to become more pervasive as sample sizes continue to increase. We also suggest that whilst analyses such as those above can unpick associations between polygenic signals impacted by confounding via geography, it would be more informative still to include such sensitivity analyses at the GWAS stage of analysis prior to subsequent analysis using derived PGI.

We suggest that genetic epidemiologists should routinely check for the presence of confounding via geography, for instance via the Hausman test for within- and between-estimate equivalence (Hausman, 1978). Adopting such sensitivity analyses may prevent future researchers from assuming the conditional exchangeability of individuals living within central London with individuals in the Peak District (Figure 5).

Researchers may even go further and explicitly interrogate the causal make up of contextual signals themselves. For instance, novel methods in multivariable mendelian randomisation (MVMR) (Sanderson et al., 2019) present an opportunity to adjust for SNPs associated with area-means of a trait in an individual-level analysis, to ensure ostensibly individual genetic signals are not a consequence of geographically structured confounds. Such work will help ensure that researchers investigating the genetic signals of complex behavioural and social traits are not unintentionally misattributing genetic causes to upstream origins of such traits.

## Methods

### Cohort Description

The UK Biobank is a large population-based prospective health research resource of 503,317 participants, aged between 38 and 73 years at recruitment, recruited between 2006 and 2010, from across the UK (Sudlow et al., 2015). Baseline assessment took place at 22 assessment centres in the UK to enable recruitment from a range of locations. Further information can be found on the UK Biobank website (www.ukbiobank.ac.uk). UK Biobank received ethical approval from the Research Ethics Committee (REC reference for UK Biobank is 11/NW/0382). We excluded participants based on the latest withdrawal lists at the time of data extraction (25/04/2023) for our UK Biobank project (project number: 81499).

### Outcome Derivation

Whether methodologically treated as an outcome or an exposure, greenspace is inconsistently defined in the literature (Cusack et al., 2017). Therefore, we used two measures to capture participants’ residential access to greenspace. These measures were made available to UK Biobank and were derived using exact postcodes, an UK-wide alphanumeric reference used to identify postal areas (Sarkar et al., 2015). Both measures were standardised prior to inclusion in models.

#### Percentage greenspace

The first outcome refers to the percentage of an area within a 300m radius of the participants home classified as greenspace. This is based on land use data from the Generalized Land Use Database (GLUD) (Department for Communities and Local Government, 2007) for England 2005 data at the 2001 census output area (OA) level. Census output areas were designed to have a consistent population between 100 and 625 persons (Office for National Statistics, 2022), thus their size varies considerably with population density. Each participant’s home location was allocated an area weighted mean of these data for output areas intersecting the 300m radius of a home. After exclusions and including only those with genetic data, the final sample size used in analyses with this outcome was 293,851 (see Figure 3). We refer to this measure as greenspace throughout.

#### Normalised Difference Vegetation Index

The second outcome was the mean Normalised Difference Vegetation Index (NDVI), calculated for a 500m radius of the participant’s home location. NDVI is a commonly used parameterisation of green-space access as it can be calculated from satellite imagery, rather than requiring administrative land use information (Herrera et al., 2018; Wang et al., 2019), and closely approximates subjective exposure estimates (Rhew et al., 2011). NDVI can take values between −1 and +1, with positive values indicating greener areas based on reflectance measures in Colour InfraRed (CIR) satellite data (Sarkar et al., 2015). The distribution in the UK Biobank sample was −0.50 to 0.58. After exclusions based on withdrawn consent and quality control filtering for genetic data (described below) and including only those with genetic data, the final sample size for NDVI was 209,391 (see Figure 6).

**Figure 6.**
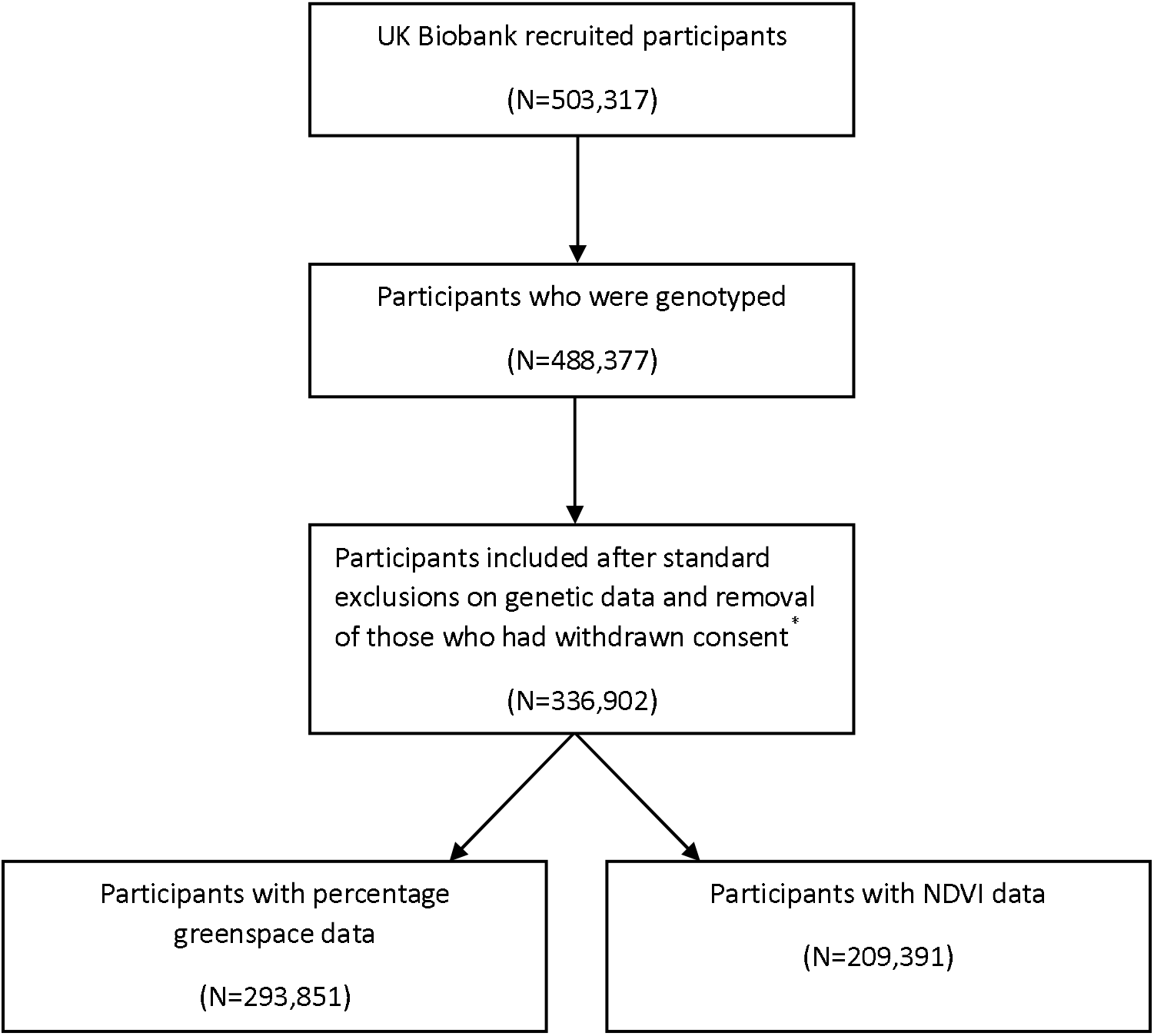
Flowchart of number of participants included in analyses (*Participants who withdrew consent by 25/04/2023), NDV I= Normalised Difference Vegetation Index.

The percentage greenspace measure was only weakly correlated with the NDVI measure (*r*=0.10). This is likely because they capture different aspects of greenspace; the NDVI measure captures greenness via the reflectance of the land surface, whereas the percentage greenspace classification captures functional greenspace, including areas of natural agricultural land, recreational areas and other areas of grassland, and excluding domestic gardens (Department for Communities and Local Government, 2007).

### Genetically proxied exposures

There were 488,377 participants with genotyped samples, of which 49,979 were genotyped using the UK BiLEVE array and 438,398 using the UK Biobank axiom array. Pre-imputation quality control, phasing and imputation have been described elsewhere (Bycroft et al., 2018). For further detail of the genetic quality controls see Supporting Information Section 2.

### Construction of polygenic indices

We constructed 12 different PGI using Plink (version 2) (Purcell et al., 2007) for each phenotype using summary statistics from genome wide association studies (GWAS) of major depressive disorder (MDD) (Wray et al., 2018), wellbeing (Okbay et al., 2016) and schizophrenia (Ripke et al., 2014). The PGI were constructed based on different p-value thresholds of the trait GWAS (0.5, 0.4, 0.3, 0.2, 0.1, 0.05, 0.01, 1×10^−3^, 1×10^−4^, 1×10^−5^, 1×10^−6^, 5×10^−8^) where available. Further details of the PGI construction are available in Supporting Information Section 3.

To further contextualise the genetic relationship observed, we examine the association between phenotypic mental health exposures and greenspace outcomes. Details of the exposures are given in Supporting Information Section 1.

#### Confounder Specification

We included age at time of initial assessment and sex in all of our analyses. We included the first 25 principal components of population structure (PCs) as confounders in all our models including PGIs. The number of PCs was chosen to reduce geographical confounding. A recent study found that adjusting for 18 PCs was a sensible correction for population structure (Sarmanova et al., 2020), but in this study we corrected for 25 PCs as this was where we saw a flattening in the variance explained by the PCs in our outcomes (Supplementary Figure S1).

#### Structural Indicators

For the multilevel specification, we assigned each observation to the administrative area containing the reported residential address. Addresses were matched on easting and northing coordinates (Office for National Statistics, 2020) to the most recently defined local administrative units. To derive a neighbourhood classification, we assigned English and Welsh addresses to Middle Layer Super Output Areas (MSOAs, average population = 7,200) and Scottish addresses to Intermediate Zones (IZ, average population = 4,200). Supplementary Figure S2 shows the spatial distribution of UK Biobank participants per MSOA/IZ and Supplementary Figure S3 gives the counts of individuals per MSOA/IZ. Data were rounded to the nearest kilometre to preserve anonymity, meaning some participants’ recorded locations fell outside of census boundaries (0.6% of participants). These participants were assigned to the nearest neighbourhood based on Euclidean distance to the census output area centre.

### Statistical Models

Analyses were conducted in R version 4.3.3 (R Core Team, 2016). We regressed both greenspace outcomes on each PGI outcome (MDD, wellbeing and schizophrenia), repeated for exposures constructed at a range of p-value thresholds. For each of these models we adjusted for sex, age and the first 25 PCs of population structure.

Multilevel models were fitted using the R package *lme4* (Bates et al., 2015). For both percentage greenspace and NDVI outcomes; across depression, wellbeing and schizophrenia PGI exposures; individuals were clustered into neighbourhoods, with the intercept allowed to vary by neighbourhood, and adjusted for sex, age, 25 PCs, and (for Mundlak models) area mean PGI as fixed effects. See Supporting Information Section 4 for algebraic detailing and interpretation of the Mundlak multilevel formulation.

For our examining of analogous phenotypic associations we specify a linear regression of greenspace outcomes on depression diagnosis, MHQ depression, wellbeing and schizophrenia, adjusted for age and sex. Results of the phenotypic associations are given in Supplementary Table S1.

## Data availability

UK Biobank data are available through a procedure described at: https://www.ukbiobank.ac.uk/enable-your-research.

## Code availability

The analysis code that forms the basis of the results presented here is available from: https://github.com/ZoeReed/Greenspace_MH_PGS_analysis.

## Funding

This work was supported in part by the UK Medical Research Council Integrative Epidemiology Unit at the University of Bristol (Grant ref: MC_UU_00032/1 and MC_UU_00032/7). GJG was supported by an ESRC Postdoctoral Fellowship and MQ Fellows Award (Grant ref: ES/T009101/1 and MQF22\22). MRM is supported by the National Institute for Health Research Bristol Biomedical Research Centre. OSPD is funded by the Alan Turing Institute under the EPSRC grant EP/N510129/1. MRM and OSPD were also supported by the National Institute for Health Research (NIHR) Biomedical Research Centre at the University Hospitals Bristol NHS Foundation Trust and the University of Bristol. TTM is funded by the ESRC (ES/W013142/1).

## Conflict of interest statement

Authors declare no conflicts of interest

## Ethics

UK Biobank received ethical approval from the Research Ethics Committee (REC reference for UK Biobank is 11/NW/0382).

## Acknowledgements

This research has been conducted using data from UK Biobank (project ID: 81499), a major biomedical database (www.ukbiobank.ac.uk). We thank the participants of UK Biobank. Quality Control filtering of the UK Biobank data was conducted by R.Mitchell, G.Hemani, T.Dudding, L.Corbin, S.Harrison, L.Paternoster as described in the published protocol (doi: 10.5523/bris.1ovaau5sxunp2cv8rcy88688v). For the purpose of open access, the author(s) has applied a Creative Commons Attribution (CC BY) licence to any Author Accepted Manuscript version arising from this submission.

## Author contributions

Conceptualization: ZER, MRM, GJG; Methodology: ZER, GJG, TTM; Formal Analysis: ZER, GJG; Resources: MRM, OSPD, GDS; Data Curation: ZER; Writing—Original Draft: ZER, GJG; Writing—Review and Editing: ZER, GJG, TTM, OSPD, GDS, MRM; Supervision: MRM, GJG; Project Administration: MRM; Funding Acquisition: MRM, GDS, OSPD

## Footnotes

1 Neighbourhoods are taken from Middle Layer Super Output Areas in England and Wales, and Intermediate Zones in Scotland, calculated from the rounded residential coordinates given for each participant in UKB.

## Notes

### Competing Interest Statement

The authors have declared no competing interest.

### Author Declarations

The UK Biobank has approval from the North West Multi-centre Research Ethics Committee as a research tissue bank. This means that researchers can operate under this approval with no need for further ethical approval, other than exceptions such as re-contact applications. This RTB approval was granted initially in 2011 and it is renewal on a five-yearly cycle: we successfully applied to renew it in 2016 and 2021. UK Biobank will apply for renewal effective in 2026.

### Summary of Updates

The manuscript has undergone internal review to ensure it is more pedagogically appropriate for the intended audience. This has involved some rewrites, and the reordering of results and methods. Some figure headings have changed, and the abstract condensed.

