## Supplementary Information for "Accounting for place: confounding via geography obscures polygenic evidence on mental health and environmental exposures in the UK Biobank"

**Supporting Information**

### 1. **Mental health phenotyping**

The UK Biobank aimed to identify determinants of human disease in middle and old age. A range of data have been collected across sociodemographic, health status, lifestyle, cognitive function, self-reported measures and physical and mental health measures. Data were collected via a number of methods, including paper and web-based questionnaires, computer assisted interviews, clinic visits and data linkage.

*Depression phenotypes:*

We included two measures of phenotypic depression. The first measure was defined using data from a combination of ICD-10 diagnoses and diagnoses reported in verbal interviews with a nurse at the assessment centre at the participants intial assessment centre visit.

For the diagnosis of depression variable, anyone who had a diagnosis based on data from a combination of ICD-10 diagnoses (ICD-10 codes used: F32, F320, F321, F322, F323, F328, F329, F33, F330, F331, F332, F333, F334, F338, F339) and diagnoses reported in verbal interviews with a nurse at the assessment centrewas coded as having depression. In UK Biobank, 14.40% of participants were coded as ever having had depression, in line with National Institute for Health and Care Excellence (NICE) estimates (14.6% lifetime prevalence).

The second measure was based on a questionnaire a sub-sample of UK Biobank participants completed as part of an on-line mental health self-assessment questionnaire issued in 2016. A variable was derived that identified those as having depression if they had probable recurrent major depression (severe or moderate) or a single probable major depression episode (Smith et al., 2013).

Anyone who had a diagnosis of depression from any of these sources was coded as having depression. . For the mental health questionnaire (MHQ) depression measure, data were available for a sub-sample of UK Biobank participants who completed questions related to depressive and manic symptoms during the final two years of recruitment (in 2016). From these data, a variable for having bipolar disorder and major depression was derived, where a participant reporting both was coded only as having the most severe condition (Smith et al., 2013). We excluded participants who had bipolar disorder type 1 or 2 and identified those as having depression if they had probable recurrent major depression (severe or moderate) or a single probable major depression episode. We refer to this as the mental health questionnaire (MHQ) depression measure.

Anyone without a diagnosis indicated in these data sources was coded as not having depression

*Schizophrenia phenotype:*

As with the depression diagnosis variable, we used data from a combination of both ICD-10 diagnoses (ICD-10 codes: F20, F200, F201, F202, F203, F204, F205, F206, F208, F209) and diagnoses reported in verbal interviews with a nurse at the assessment centre to code whether someone had a diagnosis of schizophrenia. Participants with a diagnosis of schizophrenia from any of these sources were coded as having schizophrenia. Participants without a diagnosis indicated in these data sources were coded as not having schizophrenia.

In UK Biobank, 0.22% of participants were coded as having schizophrenia, compared to general population 7.2 per 1000 median lifetime risk for schizophrenia (McGrath et al., 2008). This is consistent with known selection pressures in UK Biobank, as demonstrated previously with reduced participation in individuals with greater liability to schizophrenia (Tyrrell et al., 2021).

*Wellbeing phenotype:*

As part of the on-line mental health self-assessment questionnaire issued in 2016, participants were asked the question ‘In general how happy are you?’. Participants could select from the following options: ‘Prefer not to answer’, ‘Do not know’, ‘Extremely happy’, ‘Very happy’, ‘Moderately happy’, ‘Moderately unhappy, ‘Very unhappy’ and ‘Extremely unhappy’. We excluded participants who selected either of the first two options and coded the other options numerically so that a higher number indicated a more positive response. The score had a range of 1 to 6.

### 2. **Genetic quality control information**

In summary, multiallelic SNPs and those with a MAF≤1% were removed. Phasing of genotype data was performed using a modified version of the SHAPEIT2 algorithm. The SNPs used were imputed to the Haplotype Reference Consortium (HRC) reference panel, using IMPUTE2 algorithms. A graded filtering with different imputation qualities for different MAF ranges was used (Info>0.3 for MAF>3%, Info>0.6 for MAF 1-3%, Info>0.8 for MAF 0.5-1% and Info>0.9 for MAF 0.1-0.5%), where MAF and info scores were recalculated on an in-house derived ‘European’ subset. Individuals with sex-mismatch or sex-chromosome aneuploidy were excluded (N=814). In-house quality control filtering of the UK Biobank data is described in a published protocol(Mitchell et al., 2019). We restricted the sample to individuals of white British ancestry who self-report as “White British” and who have very similar ancestral backgrounds according to principal components analysis (N=409,703), as described by Bycroft and colleagues (Bycroft et al., 2018). Estimated kinship coefficients using the KING toolset (Manichaikul et al., 2010) identified 107,162 pairs of related individuals. An in-house algorithm was then applied to this list and preferentially removed the individuals related to the greatest number of other individuals until no related pairs remain. These individuals were excluded (N=79,450).

### 3. **Polygenic risk score construction**

The summary statistics for the MDD and wellbeing GWAS excluded UK Biobank to avoid sample overlap, and also excluded 23andMe participants. We present results across different p-value thresholds to assess the consistency of the observed effects across these different thresholds. For context, the PGI constructed at a p-value threshold of 0.05, the depression PGI explained 0.4% of the variance in having a diagnosis of depression in the UK Biobank, the wellbeing PGI explained 0.3% of the variance in wellbeing, and the schizophrenia PGI explained 2.6% of the variance in having a diagnosis of schizophrenia. SNPs from the GWAS and UK Biobank were harmonised, aligning the effect estimates and alleles. SNPs were clumped used the European subsample of the 1000 genomes project, with R^2^ < 0.25 and a window of 500 kb. The PGI were created by multiplying the number of effect alleles for each participant in UK Biobank by the effect estimate of the SNP from the relevant GWAS, then summing across all SNPs associated with each trait. PGI were z-standardised; results should be interpreted as per standard deviation (SD) increase in score.

### 4. **Random effect Mundlak Multilevel models (MLM)**

In our study we fit multilevel (or hierarchical/mixed) models (MLM) to estimate within-area effects of the polygenic indicator (PGI) exposure, whilst accounting for between-area differences in both the outcome (greenspace) and PGI exposure. A simplified traditional multilevel specification is given below:

Equation 1: Generalised Mundlak Multilevel Formulation

$$Y_{ij}= \beta_{0j}+\beta_{1W}x_{1ij}+\beta_{2C}\bar{x}_{1j}+ \Sigma_{1}^{M}\beta_{M}x_{Mij}+u_{0j}+e_{0ij}$$

$$\beta_{0j}= \beta_{0}+u_{0j}$$

$$\left[ u_{0j} \right]\sim N\left( 0, \sigma_{u0}^{2} \right)$$

$$\left[ e_{0ij} \right]\sim N\left( 0, \sigma_{e0}^{2} \right)$$

Here $Y_{ij}$ denotes the greenspace outcome for individual *i* in neighbourhood *j*. Similarly, $x_{1ij}$ denotes the mental health PGI for individual *i* in neighbourhood *j,* and ${\Sigma_{1}^{M}x}_{Mij}$ denotes measures of individual covariates for the same individual *i* in neighbourhood *j.* $u_{0j}$ denotes the error term for each neighbourhood *j* around the global mean, while $e_{0ij}$ denotes the individual error term from the neighbourhood mean.

The multilevel specification allows us to model the greenspace outcome for individuals in a given neighbourhood, relative to the average greenspace in that neighbourhood. This is because the intercept term for neighbourhood *j*, $\beta_{0j}$, is constituted of two estimated parameters. $\beta_{0}$ gives the overall intercept, the average greenspace value for a typical individual in a typical neighbourhood, where all other covariates are zero. $u_{0j}$ gives the differential greenspace associated with living in neighbourhood *j*. These neighbourhood differentials are assumed to come from a normal distribution, with mean 0 and variance $\sigma_{u0}^{2}$. This variance represents the variation in greenspace outcomes between neighbourhoods.

By convention we have given the $\beta$ estimates associated with $x_{1}$ subscripts indicating the within- and contextual- estimates (Bell et al., 2019).

As such, $\beta_{1W}$ estimates the average greenspace change for a unit increase in mental health PGI, *within the average neighbourhood*, having taken account of between-neighbourhood differences via the random-intercept specification. $\beta_{1C}$ gives the contextual effect of a level-1 individual moving from one neighbourhood to another where the mean PGI is 1 unit higher, holding the characteristics of the individual constant.

If we were instead interested in the effect of changing the level of $\bar{x}_{1j}$ whilst not requiring that the effect of $x_{1ij}$ remains the same, we could instead subtract the mean $\bar{x}_{1j}$ from our $x_{1ij}$ estimate, which would change the effect of the $\bar{x}_{1j}$ term to capture a between-area estimate ($\beta_{2B})$, where individual characteristics were not held constant.

These models are necessarily equivalent, however, where $\beta_{1W}+ \beta_{2C}= \beta_{2B}$, (where such that the between area effect can be calculated from the anomaly of the within and contextual effect estimates. It is exactly the $\beta_{2C}$ term that is assumed to be zero in a single level model, as the within- and between- effect estmiates are necessarily assumed to be exactly equal.

$\beta_{M}$ represents the coefficients for remaining individual-level covariates in the adjustment set.

Finally, the residual unexplained, individual-level variation is given by $e_{0ij}$, and is similarly assumed to come from a normal distribution with mean 0 and variance $\sigma_{e0}^{2}$. From the variance components, $\sigma_{u0}^{2}$ and $\sigma_{e0}^{2}$, we can calculate the Variance Partitioning Coefficient (VPC), which assesses the proportion of total variation in outcome which is accounted for by a specific level in the model. In the two-level model formulation, the VPC is exactly equal to the intra-class correlation coefficient (ICC). The ICC gives the expected correlation between randomly selected pairs of lower level units from within the same higher level unit. For instance, we could calculate the Level-2 VPC for greenspace as follows:

Equation 2: Level 2 Variance Partitioning Formula

$$Level 2 VPC for Y_{\mathrm{ij}}=\frac{\sigma_{u0}^{2}}{\sigma_{u0}^{2}+\sigma_{e0}^{2}}$$

In our example, this would tell us what proportion of the total unexplained variation in greenspace is accounted for by the neighbourhood in which someone lives.

Similarly, in the ICC interpretation, it tells us the expected correlation between greenspace outcomes for pairs of individuals drawn from the same neighbourhood. Higher VPC or ICC values imply greater similarity in greenspace amongst individuals living in the same neighbourhood. This is particularly important for our investigation, as we know that greenspace is likely to be very similar amongst individuals who live within the same neighbourhood due to its derivation from environmental information around a residential location.

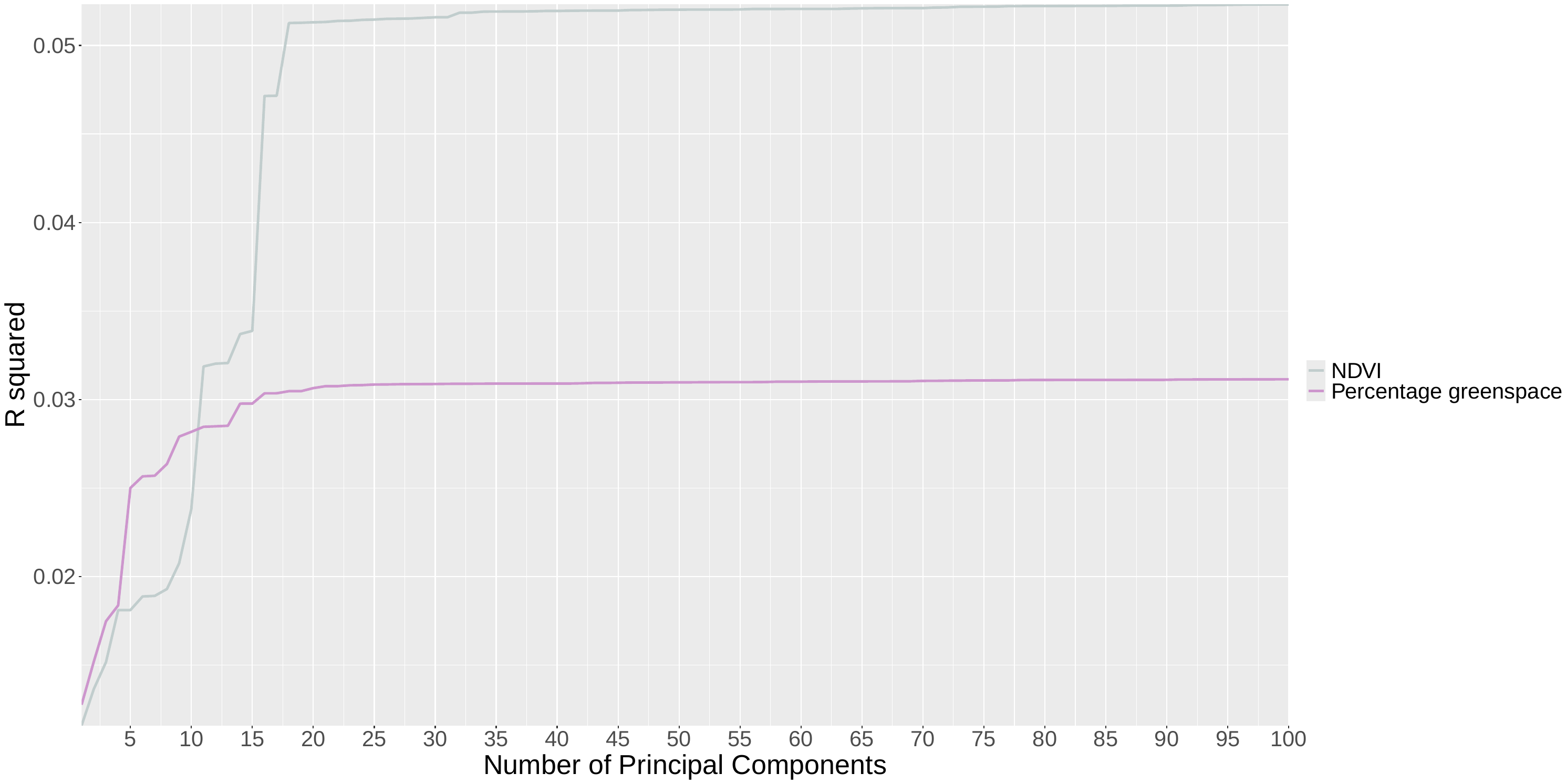

***Supplementary Figure S1.*** *Modelled R-squared values with increasing numbers of genetic principal components for Normalised Difference Vegetation Index (NDVI) and Percentage Greenspace (derived from land-use).*

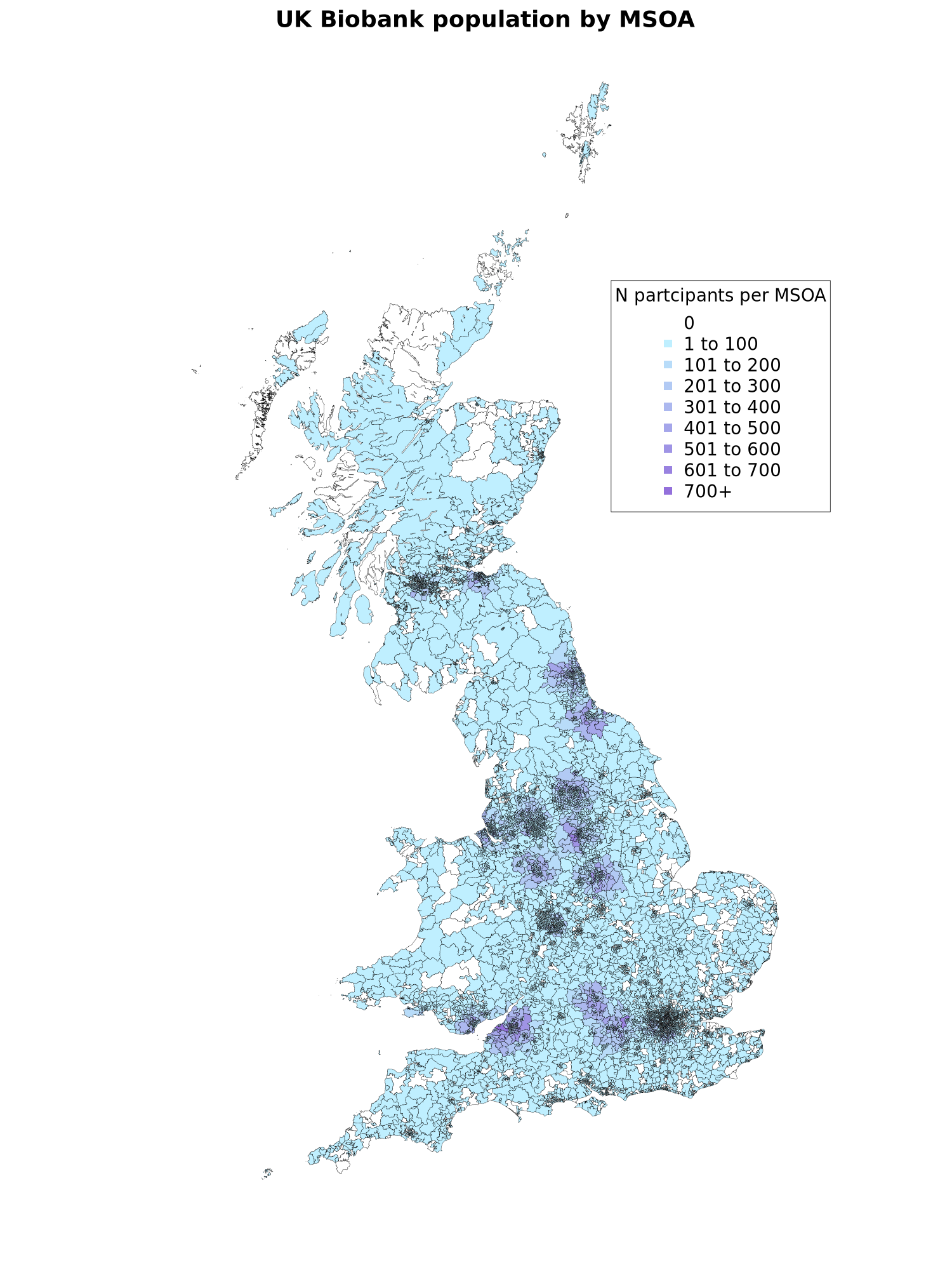

***Supplementary Figure S2.*** *Spatial distribution of number of participants per MSOA (or equivalent) in UK Biobank. Spatial polygons represent Middle Layer Super Output Areas (MSOAs) for England and Wales and Intermediate Zones (IZs) for Scotland.*

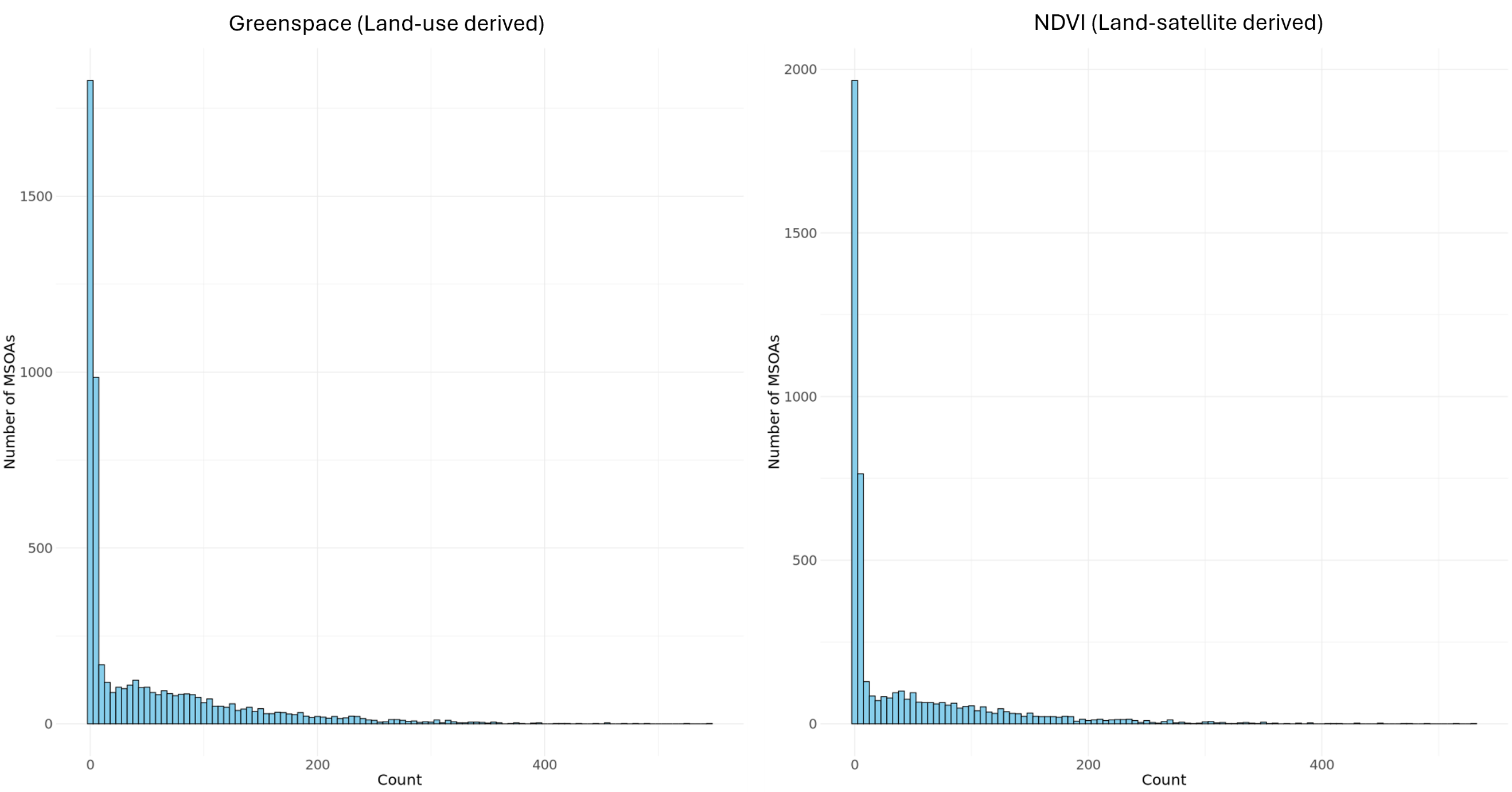

***Supplementary Figure S3.*** *Histograms of MSOA populations for Greenspace (land-use derived) outcome and Normalised Difference Vegetation Index (NDVI, Land-satellite derived) outcome. Total MSOAs for Greenspace analyses = 5728, total MSOAs for NDVI analyses = 4896). Y-axis gives frequency of observed count, and X-axis gives number of observed individuals in an MSOA.*

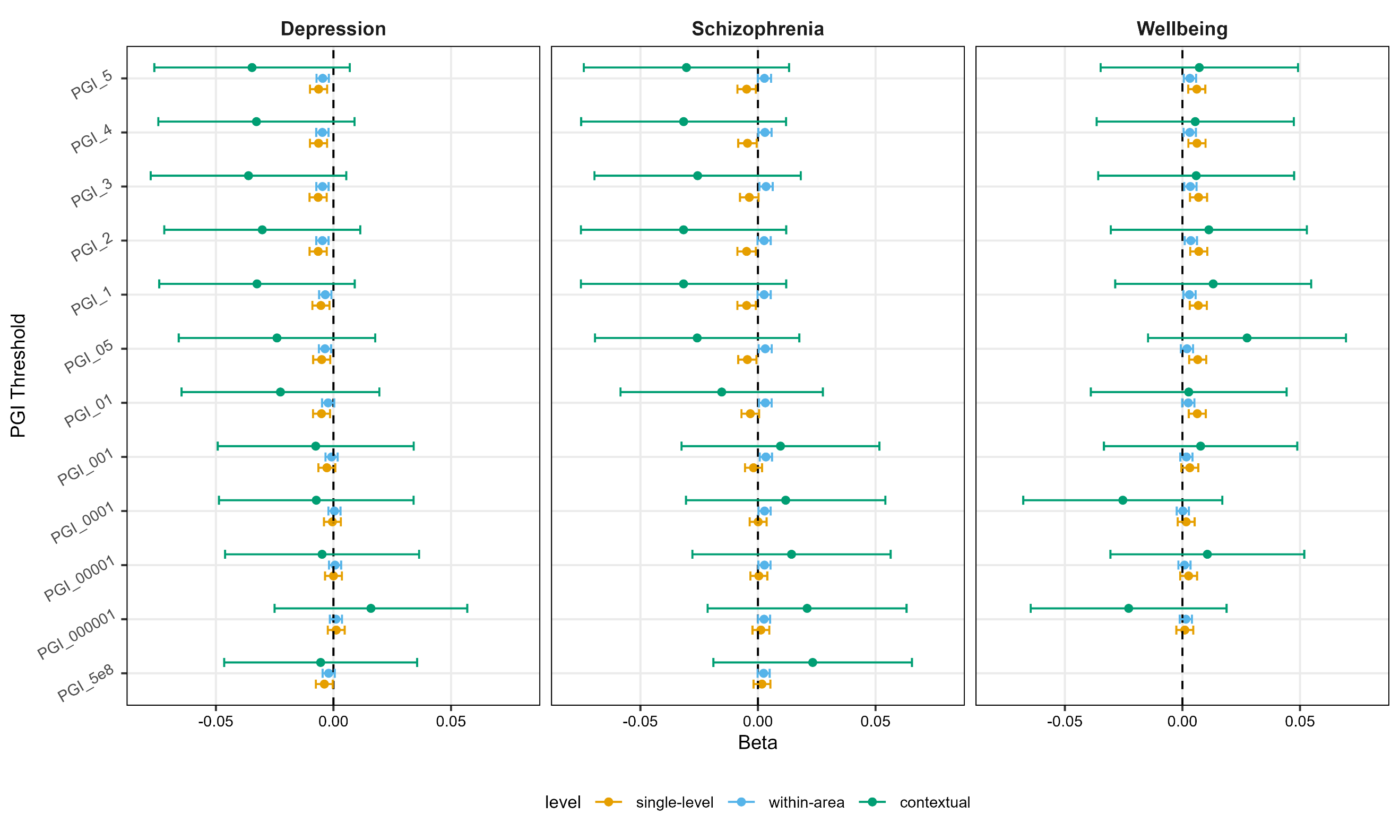

***Supplementary Figure S4*.** *Polygenic mental health exposures and Greenspace (landuse percentage) outcome, with contextual effects displayed alongside within-area, and single-level estimates. PGI = Polygenic Index. PGI_5 through PGI_5e-8 denote polygenic indices constructed with SNP selection filtered at p<0.5, p<0.4, p<0.3, p<0.2, p<0.1, p<0.05, p<0.01, p<0.001, p<0.0001, p<0.00001, p<0.000001 and p< 5x10^-8^ in descending order down the Y axis.*

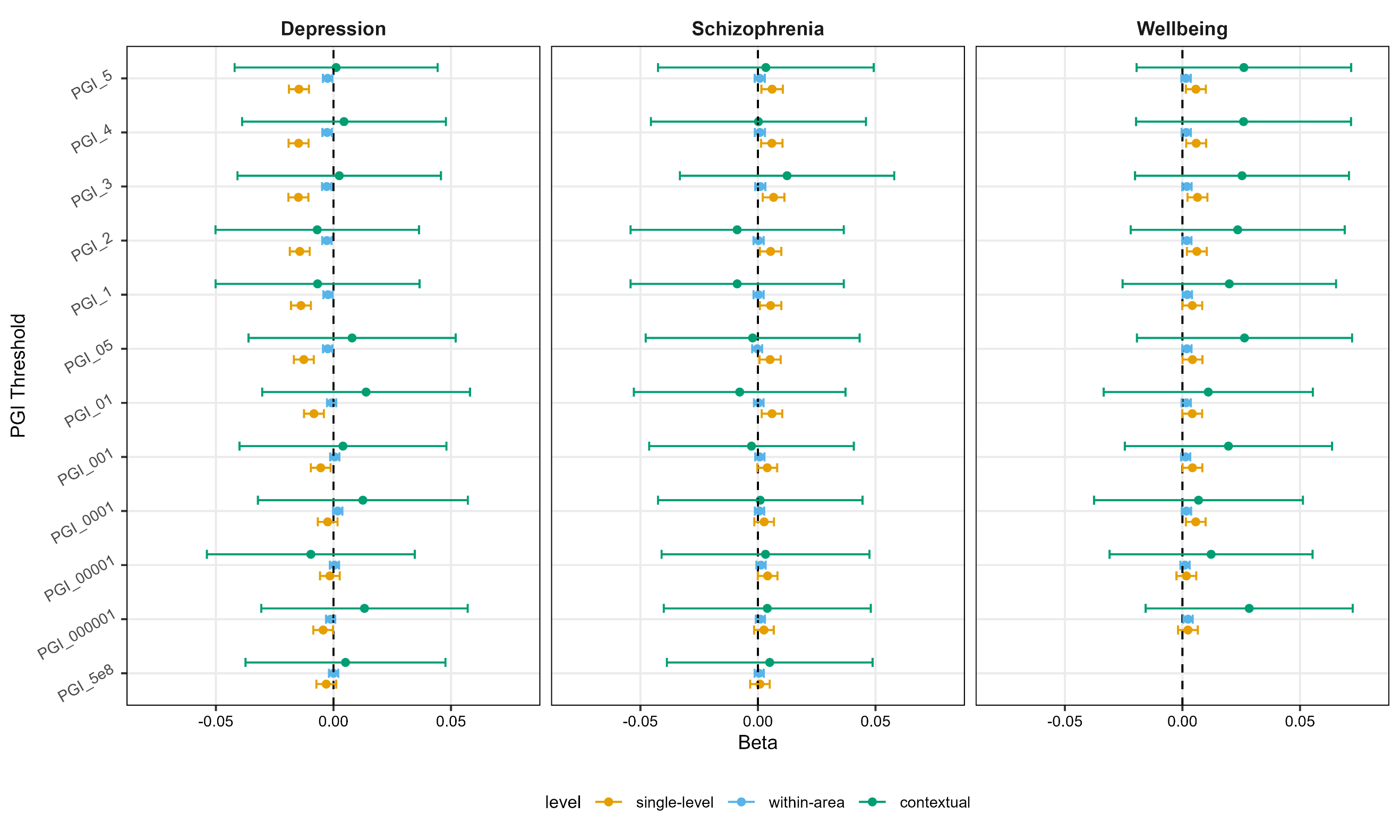

***Supplementary Figure S5.*** *Polygenic mental health exposures and NDVI (Normalised Difference Vegetation Index) outcome, with contextual effects displayed alongside within-area, and single-level estimates. PGI = Polygenic Index. PGI_5 through PGI_5e-8 denote polygenic indices constructed with SNP selection filtered at p<0.5, p<0.4, p<0.3, p<0.2, p<0.1, p<0.05, p<0.01, p<0.001, p<0.0001, p<0.00001, p<0.000001 and p< 5x10^-8^ in descending order down the Y axis.*

***Supplementary Table S1.*** *Associations between depression, wellbeing and schizophrenia phenotypes and Normalised Difference Vegetation Index and percentage greenspace from linear regression models*

| Mental health phenotype | Greenspace measure | N | Beta* | 2.5% CI | 97.5% CI | P |
| --- | --- | --- | --- | --- | --- | --- |
| *Depression diagnosis* | Percentage greenspace | 293,851 | -0.07 | -0.08 | -0.06 | 4.59x10^-37^ |
|  | NDVI | 209,391 | -0.005 | -0.02 | 0.007 | 0.44 |
| *MHQ depression* | Percentage greenspace | 77,335 | -0.03 | -0.05 | -0.01 | 1.69x10^-04^ |
|  | NDVI | 54,061 | -0.02 | -0.04 | -0.004 | 0.01 |
| *Wellbeing* | Percentage greenspace | 97,076 | 0.06 | 0.05 | 0.07 | 9.65x10^-42^ |
|  | NDVI | 67,737 | -0.01 | -0.02 | -0.0003 | 0.04 |
| *Schizophrenia* | Percentage greenspace | 293,851 | -0.32 | -0.40 | -0.25 | 7.35x10^-17^ |
|  | NDVI | 209,391 | 0.04 | -0.05 | 0.13 | 0.40 |

*CI=Confidence Interval, NDVI=Normalised Difference Vegetation Index, MHQ=Mental Health Questionnaire. *Beta reflects the NDVI or percentage greenspace SD change predicted by depression or schizophrenia diagnosis or every unit increase in the wellbeing score*

***Supplementary Table S2.*** *Single level effect estimates of percentage greenspace regressed on polygenic scores for depression, wellbeing and schizophrenia from linear (single-level) models for PGI at all p-value thresholds (N=293,851)*

| Phenotype | Polygenic indicator p-value threshold | Beta | 2.5% CI | 97.5% CI | P |
| --- | --- | --- | --- | --- | --- |
| Depression | 0.5 | -0.006 | -0.01 | -0.003 | 0.0007 |
|  | 0.4 | -0.006 | -0.01 | -0.003 | 0.0006 |
|  | 0.3 | -0.007 | -0.01 | -0.003 | 0.0005 |
|  | 0.2 | -0.007 | -0.01 | -0.003 | 0.0005 |
|  | 0.1 | -0.005 | -0.009 | -0.002 | 0.004 |
|  | 0.05 | -0.005 | -0.009 | -0.001 | 0.007 |
|  | 0.01 | -0.005 | -0.009 | -0.001 | 0.006 |
|  | 1x10-03 | -0.003 | -0.006 | 0.0008 | 0.12 |
|  | 1x10-04 | -0.0004 | -0.004 | 0.003 | 0.81 |
|  | 1x10-05 | 1.00E-05 | -0.004 | 0.004 | 1.0 |
|  | 1x10-06 | 0.001 | -0.002 | 0.005 | 0.52 |
|  | 5x10-08 | -0.004 | -0.007 | -0.0003 | 0.04 |
| Schizophrenia | 0.5 | -0.005 | -0.009 | -0.0009 | 0.02 |
|  | 0.4 | -0.004 | -0.008 | -0.0005 | 0.03 |
|  | 0.3 | -0.004 | -0.008 | 0.0003 | 0.07 |
|  | 0.2 | -0.005 | -0.009 | -0.0009 | 0.02 |
|  | 0.1 | -0.005 | -0.009 | -0.0009 | 0.02 |
|  | 0.05 | -0.005 | -0.008 | -0.0006 | 0.02 |
|  | 0.01 | -0.003 | -0.007 | 0.0005 | 0.09 |
|  | 1x10-03 | -0.002 | -0.006 | 0.002 | 0.31 |
|  | 1x10-04 | 8.00E-05 | -0.004 | 0.004 | 0.96 |
|  | 1x10-05 | 0.0004 | -0.003 | 0.004 | 0.84 |
|  | 1x10-06 | 0.001 | -0.002 | 0.005 | 0.5 |
|  | 5x10-08 | 0.002 | -0.002 | 0.005 | 0.35 |
| Wellbeing | 0.5 | 0.006 | 0.002 | 0.01 | 0.001 |
|  | 0.4 | 0.006 | 0.003 | 0.01 | 0.0009 |
|  | 0.3 | 0.007 | 0.003 | 0.01 | 0.0002 |
|  | 0.2 | 0.007 | 0.003 | 0.01 | 0.0002 |
|  | 0.1 | 0.007 | 0.003 | 0.01 | 0.0003 |
|  | 0.05 | 0.006 | 0.003 | 0.01 | 0.0005 |
|  | 0.01 | 0.006 | 0.003 | 0.01 | 0.0007 |
|  | 1x10-03 | 0.003 | -0.0005 | 0.007 | 0.09 |
|  | 1x10-04 | 0.002 | -0.002 | 0.005 | 0.38 |
|  | 1x10-05 | 0.003 | -0.0009 | 0.006 | 0.15 |
|  | 1x10-06 | 0.001 | -0.003 | 0.005 | 0.58 |

*CI=Confidence Interval. *Beta reflects the percentage greenspace SD change predicted by an SD increase in the polygenic score for the average individual*

***Supplementary Table S3.*** *Single level effect estimates of NDVI regressed on polygenic scores for depression, wellbeing and schizophrenia from linear (single-level) models for PGI at all p-value thresholds (N=209,391)*

| Phenotype | Polygenic indicator p-value threshold | Beta* | 2.5% CI | 97.5% CI | P |
| --- | --- | --- | --- | --- | --- |
| *Depression* | 0.5 | -0.01 | -0.02 | -0.01 | 1.32x10-11 |
|  | 0.4 | -0.01 | -0.02 | -0.01 | 8.30x10-12 |
|  | 0.3 | -0.01 | -0.02 | -0.01 | 5.77x10-12 |
|  | 0.2 | -0.01 | -0.02 | -0.01 | 3.65x10-11 |
|  | 0.1 | -0.01 | -0.02 | -0.01 | 1.53x10-10 |
|  | 0.05 | -0.01 | -0.02 | -0.008 | 5.14x10-09 |
|  | 0.01 | -0.008 | -0.01 | -0.004 | 0.0001 |
|  | 1x10-03 | -0.005 | -0.01 | -0.001 | 0.01 |
|  | 1x10-04 | -0.002 | -0.007 | 0.002 | 0.25 |
|  | 1x10-05 | -0.002 | -0.006 | 0.002 | 0.47 |
|  | 1x10-06 | -0.004 | -0.009 | -0.0002 | 0.04 |
|  | 5x10-08 | -0.003 | -0.007 | 0.001 | 0.15 |
| *Schizophrenia* | 0.5 | 0.006 | 0.001 | 0.01 | 0.01 |
|  | 0.4 | 0.006 | 0.001 | 0.01 | 0.01 |
|  | 0.3 | 0.007 | 0.002 | 0.01 | 0.005 |
|  | 0.2 | 0.005 | 0.0008 | 0.01 | 0.02 |
|  | 0.1 | 0.005 | 0.0008 | 0.01 | 0.02 |
|  | 0.05 | 0.005 | 0.0006 | 0.01 | 0.03 |
|  | 0.01 | 0.006 | 0.002 | 0.01 | 0.007 |
|  | 1x10-03 | 0.004 | -0.0003 | 0.008 | 0.07 |
|  | 1x10-04 | 0.003 | -0.002 | 0.007 | 0.22 |
|  | 1x10-05 | 0.004 | -6.00E-05 | 0.008 | 0.05 |
|  | 1x10-06 | 0.003 | -0.002 | 0.007 | 0.23 |
|  | 5x10-08 | 0.0009 | -0.003 | 0.005 | 0.69 |
| *Wellbeing* | 0.5 | 0.006 | 0.001 | 0.01 | 0.009 |
|  | 0.4 | 0.006 | 0.002 | 0.01 | 0.007 |
|  | 0.3 | 0.006 | 0.002 | 0.01 | 0.003 |
|  | 0.2 | 0.006 | 0.002 | 0.01 | 0.005 |
|  | 0.1 | 0.004 | -6.00E-05 | 0.008 | 0.05 |
|  | 0.05 | 0.004 | 4.00E-05 | 0.009 | 0.05 |
|  | 0.01 | 0.004 | -7.00E-05 | 0.008 | 0.05 |
|  | 1x10-03 | 0.004 | 6.00E-05 | 0.008 | 0.05 |
|  | 1x10-04 | 0.006 | 0.002 | 0.01 | 0.008 |
|  | 1x10-05 | 0.002 | -0.003 | 0.006 | 0.43 |
|  | 1x10-06 | 0.002 | -0.002 | 0.007 | 0.27 |

*CI=Confidence Interval. *Beta reflects the NDVI SD change predicted by an SD increase in the polygenic score for the average individual*

***Supplementary Table S4.*** *Within-area effect estimates of percentage greenspace regressed on polygenic scores for depression, wellbeing and schizophrenia for all p-value thresholds (N=293,851).*

| Phenotype | Polygenic indicator p-value threshold | Beta* | 2.5% CI | 97.5% CI | P |
| --- | --- | --- | --- | --- | --- |
| Depression | 0.5 | -0.005 | -0.007 | -0.002 | 0.0006 |
|  | 0.4 | -0.005 | -0.007 | -0.002 | 0.0004 |
|  | 0.3 | -0.005 | -0.007 | -0.002 | 0.0005 |
|  | 0.2 | -0.005 | -0.007 | -0.002 | 0.0004 |
|  | 0.1 | -0.003 | -0.006 | -0.0009 | 0.009 |
|  | 0.05 | -0.004 | -0.006 | -0.001 | 0.007 |
|  | 0.01 | -0.002 | -0.005 | 0.0003 | 0.09 |
|  | 1x10-03 | -0.0008 | -0.003 | 0.002 | 0.55 |
|  | 1x10-04 | 0.0004 | -0.002 | 0.003 | 0.75 |
|  | 1x10-05 | 0.0007 | -0.002 | 0.003 | 0.61 |
|  | 1x10-06 | 0.001 | -0.002 | 0.004 | 0.42 |
|  | 5x10-08 | -0.002 | -0.005 | 0.0005 | 0.12 |
| Schizophrenia | 0.5 | 0.003 | -6.00E-05 | 0.006 | 0.05 |
|  | 0.4 | 0.003 | 0.0002 | 0.006 | 0.04 |
|  | 0.3 | 0.003 | 0.0006 | 0.006 | 0.02 |
|  | 0.2 | 0.003 | -0.0002 | 0.005 | 0.07 |
|  | 0.1 | 0.003 | -0.0002 | 0.005 | 0.07 |
|  | 0.05 | 0.003 | 0.0003 | 0.006 | 0.03 |
|  | 0.01 | 0.003 | 0.0005 | 0.006 | 0.02 |
|  | 1x10-03 | 0.003 | 0.0008 | 0.006 | 0.01 |
|  | 1x10-04 | 0.003 | 0.0002 | 0.005 | 0.03 |
|  | 1x10-05 | 0.003 | 0.0002 | 0.005 | 0.03 |
|  | 1x10-06 | 0.003 | 3.00E-05 | 0.005 | 0.05 |
|  | 5x10-08 | 0.002 | -0.0001 | 0.005 | 0.06 |
| Wellbeing | 0.5 | 0.003 | 0.0006 | 0.006 | 0.02 |
|  | 0.4 | 0.003 | 0.0005 | 0.006 | 0.02 |
|  | 0.3 | 0.003 | 0.0008 | 0.006 | 0.01 |
|  | 0.2 | 0.004 | 0.001 | 0.006 | 0.007 |
|  | 0.1 | 0.003 | 0.0004 | 0.006 | 0.02 |
|  | 0.05 | 0.002 | -0.0007 | 0.005 | 0.15 |
|  | 0.01 | 0.002 | -0.0001 | 0.005 | 0.06 |
|  | 1x10-03 | 0.002 | -0.001 | 0.004 | 0.2 |
|  | 1x10-04 | 0.0002 | -0.002 | 0.003 | 0.9 |
|  | 1x10-05 | 0.0009 | -0.002 | 0.003 | 0.51 |
|  | 1x10-06 | 0.001 | -0.001 | 0.004 | 0.27 |

*CI=Confidence Interval, MSOA = Middle Layer Super Output Area or intermediate zone. *Beta reflects the percentage greenspace SD change predicted by an SD increase in the polygenic score, for the average individual within a typical MSOA. Estimand of* $\beta_{1W}$ *in Supplementary Section 4, Equation 1.*

***Supplementary Table S5.*** *Within-area effect estimates of NDVI regressed on polygenic scores for depression, wellbeing and schizophrenia for all p-value thresholds (N=209,391), taken from Mundlak formulated multilevel models.*

| Phenotype | Polygenic score p-value threshold | Beta* | 2.5% CI | 97.5% CI | P |
| --- | --- | --- | --- | --- | --- |
| Depression | 0.5 | -0.002 | -0.004 | -5.00E-04 | 0.01 |
|  | 0.4 | -0.003 | -0.005 | -7.00E-04 | 0.008 |
|  | 0.3 | -0.003 | -0.005 | -9.00E-04 | 0.005 |
|  | 0.2 | -0.003 | -0.005 | -8.00E-04 | 0.006 |
|  | 0.1 | -0.002 | -0.004 | -3.00E-04 | 0.02 |
|  | 0.05 | -0.002 | -0.004 | -5.00E-04 | 0.02 |
|  | 0.01 | -0.001 | -0.003 | 0.001 | 0.43 |
|  | 1x10-03 | 0.001 | -0.001 | 0.003 | 0.57 |
|  | 1x10-04 | 0.002 | -1.00E-04 | 0.004 | 0.07 |
|  | 1x10-05 | 0.000 | -0.002 | 0.002 | 0.68 |
|  | 1x10-06 | -0.001 | -0.003 | 8.00E-04 | 0.23 |
|  | 5x10-08 | 0.000 | -0.002 | 0.002 | 0.95 |
| Schizophrenia | 0.5 | 0.001 | -0.001 | 0.003 | 0.48 |
|  | 0.4 | 0.001 | -0.001 | 0.003 | 0.44 |
|  | 0.3 | 0.001 | -0.001 | 0.003 | 0.37 |
|  | 0.2 | 0.000 | -0.002 | 0.002 | 0.81 |
|  | 0.1 | 0.000 | -0.002 | 0.002 | 0.81 |
|  | 0.05 | 0.000 | -0.002 | 0.002 | 0.76 |
|  | 0.01 | 0.000 | -0.002 | 0.002 | 0.75 |
|  | 1x10-03 | 0.001 | -0.001 | 0.003 | 0.43 |
|  | 1x10-04 | 0.001 | -0.001 | 0.003 | 0.51 |
|  | 1x10-05 | 0.001 | -6.00E-04 | 0.003 | 0.19 |
|  | 1x10-06 | 0.001 | -0.001 | 0.003 | 0.31 |
|  | 5x10-08 | 0.001 | -0.001 | 0.002 | 0.61 |
| Wellbeing | 0.5 | 0.002 | -5.00E-04 | 0.004 | 0.13 |
|  | 0.4 | 0.002 | -4.00E-04 | 0.004 | 0.12 |
|  | 0.3 | 0.002 | -8.00E-05 | 0.004 | 0.06 |
|  | 0.2 | 0.002 | -1.00E-04 | 0.004 | 0.06 |
|  | 0.1 | 0.002 | 1.00E-04 | 0.004 | 0.04 |
|  | 0.05 | 0.002 | -9.00E-05 | 0.004 | 0.06 |
|  | 0.01 | 0.002 | -4.00E-04 | 0.004 | 0.12 |
|  | 1x10-03 | 0.001 | -6.00E-04 | 0.003 | 0.17 |
|  | 1x10-04 | 0.002 | -3.00E-04 | 0.004 | 0.09 |
|  | 1x10-05 | 0.001 | -9.00E-04 | 0.003 | 0.28 |
|  | 1x10-06 | 0.002 | 4.00E-04 | 0.004 | 0.02 |

*CI=Confidence Interval, MSOA = Middle Layer Super Output Area or intermediate zone. *Beta reflects the NDVI SD change predicted by an SD increase in the polygenic score, for the average individual* within a typical MSOA. *Estimand of* $\beta_{1W}$ *in Supplementary Section 4, Equation 1.*

***Supplementary Table S6.*** *Contextual effect estimates of percentage greenspace regressed on polygenic scores for depression, wellbeing and schizophrenia for all p-value thresholds (N=293,851), taken from Mundlak formulated multilevel models.*

| Phenotype | Polygenic score p-value threshold | Beta* | 2.5% CI | 97.5% CI | P |
| --- | --- | --- | --- | --- | --- |
| Depression | 0.5 | -0.03 | -0.08 | 0.007 | 0.1 |
|  | 0.4 | -0.03 | -0.07 | 0.009 | 0.12 |
|  | 0.3 | -0.04 | -0.08 | 0.005 | 0.09 |
|  | 0.2 | -0.03 | -0.07 | 0.01 | 0.15 |
|  | 0.1 | -0.03 | -0.07 | 0.009 | 0.13 |
|  | 0.05 | -0.02 | -0.07 | 0.02 | 0.26 |
|  | 0.01 | -0.02 | -0.06 | 0.02 | 0.29 |
|  | 1x10-03 | -0.008 | -0.05 | 0.03 | 0.72 |
|  | 1x10-04 | -0.007 | -0.05 | 0.03 | 0.73 |
|  | 1x10-05 | -0.005 | -0.05 | 0.04 | 0.82 |
|  | 1x10-06 | 0.02 | -0.03 | 0.06 | 0.45 |
|  | 5x10-08 | -0.005 | -0.05 | 0.04 | 0.79 |
| Wellbeing | 0.5 | 0.007 | -0.03 | 0.05 | 0.74 |
|  | 0.4 | 0.005 | -0.04 | 0.05 | 0.8 |
|  | 0.3 | 0.006 | -0.04 | 0.05 | 0.78 |
|  | 0.2 | 0.01 | -0.03 | 0.05 | 0.6 |
|  | 0.1 | 0.01 | -0.03 | 0.05 | 0.54 |
|  | 0.05 | 0.03 | -0.01 | 0.07 | 0.2 |
|  | 0.01 | 0.003 | -0.04 | 0.04 | 0.9 |
|  | 1x10-03 | 0.008 | -0.03 | 0.05 | 0.71 |
|  | 1x10-04 | -0.03 | -0.07 | 0.02 | 0.24 |
|  | 1x10-05 | 0.01 | -0.03 | 0.05 | 0.61 |
|  | 1x10-06 | -0.02 | -0.06 | 0.02 | 0.28 |
| Schizophrenia | 0.5 | -0.03 | -0.07 | 0.01 | 0.17 |
|  | 0.4 | -0.03 | -0.08 | 0.01 | 0.16 |
|  | 0.3 | -0.03 | -0.07 | 0.02 | 0.25 |
|  | 0.2 | -0.03 | -0.08 | 0.01 | 0.16 |
|  | 0.1 | -0.03 | -0.08 | 0.01 | 0.16 |
|  | 0.05 | -0.03 | -0.07 | 0.02 | 0.24 |
|  | 0.01 | -0.02 | -0.06 | 0.03 | 0.48 |
|  | 1x10-03 | 0.01 | -0.03 | 0.05 | 0.66 |
|  | 1x10-04 | 0.01 | -0.03 | 0.05 | 0.59 |
|  | 1x10-05 | 0.01 | -0.03 | 0.06 | 0.51 |
|  | 1x10-06 | 0.02 | -0.02 | 0.06 | 0.33 |
|  | 5x10-08 | 0.02 | -0.02 | 0.07 | 0.28 |

*CI=Confidence Interval, MSOA = Middle Layer Super Output Area or intermediate zone. *Beta reflects the percentage greenspace SD change predicted by an SD increase in the polygenic score for the cluster mean, holding individual level polygenic score constant. Estimand of* $\beta_{1C}$ *in Supplementary Section 4, Equation 1.*

***Supplementary Table S7.*** *Contextual effect estimates of NDVI regressed on polygenic scores for depression, wellbeing and schizophrenia for all p-value thresholds (N=209,391), taken from Mundlak formulated multilevel models.*

| Phenotype | Polygenic score p-value threshold | Beta* | 2.5% CI | 97.5% CI | P |
| --- | --- | --- | --- | --- | --- |
| *Depression* | 0.5 | 0.001 | -0.04 | 0.04 | 0.96 |
|  | 0.4 | 0.004 | -0.04 | 0.05 | 0.84 |
|  | 0.3 | 0.002 | -0.04 | 0.05 | 0.91 |
|  | 0.2 | -0.007 | -0.05 | 0.04 | 0.75 |
|  | 0.1 | -0.007 | -0.05 | 0.04 | 0.76 |
|  | 0.05 | 0.008 | -0.04 | 0.05 | 0.72 |
|  | 0.01 | 0.01 | -0.03 | 0.06 | 0.54 |
|  | 1x10-03 | 0.004 | -0.04 | 0.05 | 0.86 |
|  | 1x10-04 | 0.01 | -0.03 | 0.06 | 0.58 |
|  | 1x10-05 | -0.01 | -0.05 | 0.03 | 0.67 |
|  | 1x10-06 | 0.01 | -0.03 | 0.06 | 0.56 |
|  | 5x10-08 | 0.005 | -0.04 | 0.05 | 0.81 |
| *Schizophrenia* | 0.5 | 0.003 | -0.04 | 0.05 | 0.86 |
|  | 0.4 | 2.00E-04 | -0.05 | 0.05 | 0.99 |
|  | 0.3 | 0.01 | -0.03 | 0.06 | 0.59 |
|  | 0.2 | -0.009 | -0.05 | 0.04 | 0.7 |
|  | 0.1 | -0.009 | -0.05 | 0.04 | 0.7 |
|  | 0.05 | -0.002 | -0.05 | 0.04 | 0.92 |
|  | 0.01 | -0.008 | -0.05 | 0.04 | 0.73 |
|  | 1x10-03 | -0.003 | -0.05 | 0.04 | 0.9 |
|  | 1x10-04 | 0.001 | -0.04 | 0.05 | 0.96 |
|  | 1x10-05 | 0.003 | -0.04 | 0.05 | 0.89 |
|  | 1x10-06 | 0.004 | -0.04 | 0.05 | 0.86 |
|  | 5x10-08 | 0.005 | -0.04 | 0.05 | 0.82 |
| Wellbeing | 0.5 | 0.03 | -0.02 | 0.07 | 0.26 |
|  | 0.4 | 0.03 | -0.02 | 0.07 | 0.26 |
|  | 0.3 | 0.03 | -0.02 | 0.07 | 0.27 |
|  | 0.2 | 0.02 | -0.02 | 0.07 | 0.31 |
|  | 0.1 | 0.02 | -0.03 | 0.07 | 0.39 |
|  | 0.05 | 0.03 | -0.02 | 0.07 | 0.26 |
|  | 0.01 | 0.01 | -0.03 | 0.06 | 0.63 |
|  | 1x10-03 | 0.02 | -0.02 | 0.06 | 0.38 |
|  | 1x10-04 | 0.007 | -0.04 | 0.05 | 0.76 |
|  | 1x10-05 | 0.01 | -0.03 | 0.06 | 0.58 |
|  | 1x10-06 | 0.03 | -0.02 | 0.07 | 0.21 |

*CI=Confidence Interval, MSOA = Middle Layer Super Output Area or intermediate zone. * Beta reflects the NDVI SD change predicted by an SD increase in the polygenic score for the cluster mean, holding individual level polygenic score constant. Estimand of* $\beta_{1C}$ *in Supplementary Section 4, Equation 1.*
